# Irradiation of the Subventricular Zone and Subgranular Zone: an Atlas-based analysis on Overall Survival in High- and Low-Grade Glioma Patients

**DOI:** 10.1101/2021.08.17.21262102

**Authors:** Danique E. Bruil, Szabolcs David, Steven H.J Nagtegaal, Sophia F.A.M de Sonnaville, Joost J.C. Verhoeff

**Affiliations:** Department of Radiation Oncology, University Medical Center Utrecht, HP Q 00.3.11, PO box 85500, 3508 GA, The Netherlands; Department of Neurosurgery, University Medical Center Utrecht, HP G 03.129, PO box 85500, 3508 GA, The Netherlands

**Keywords:** Radiotherapy, Glioma, Subventricular Zone, Subgranular Zone, Overall Survival

## Abstract

**Background:** Neural stem cells in the subventricular- (SVZ) and subgranular zone (SGZ) are hypothesized to support growth of glioma. Therefore, irradiation of the SVZ and SGZ might reduce tumor growth and might improve overall survival (OS). However, it may also inhibit the repair capacity of brain tissue. The aim of this retrospective cohort study is to assess the impact of SVZ and SGZ radiotherapy doses on OS of patients with high-grade (HGG) or low-grade (LGG) glioma.

**Methods:** We included 273 glioma patients who received radiotherapy. We created an SVZ atlas, shared openly with this work, while SGZ labels were taken from the CoBRA atlas. Next, SVZ and SGZ regions were automatically delineated on T1 MR-images. Dose and OS correlations were investigated with Cox regression and Kaplan-Meier analysis.

**Results:** Cox regression analyses showed significant hazard ratios for SVZ dose (univariate: 1.029/Gy, *p*<0.001; multivariate: 1.103/Gy, *p* = 0.002) and SGZ dose (univariate: 1.023/Gy, *p*<0.001; multivariate: 1.055/Gy, *p*<0.001) in HGG patients. Kaplan-Meier analysis showed significant correlations between OS and high/low dose groups for HGG patients (SVZ: respectively 10.7 months (>30.33 Gy) vs 14.0 months (<30.33 Gy) median OS, *p* = 0.011; SGZ: respectively 10.7 months (>29.11 Gy) vs 15.5 months (<29.11 Gy) median OS, *p*<0.001). No correlations between dose and OS were not found for LGG patients.

**Conclusion:** Irradiation doses on neurogenic areas correlate negatively with OS in patients with HGG. Whether sparing of the SVZ and SGZ during radiotherapy improves OS, should be subject of prospective studies.

**Key Points:**

- Neural stem cells in the SVZ and SGZ are hypothesized to support growth of glioma.
- Higher radiation doses on the SVZ and SGZ correlate with lower OS in HGG patients.
- Avoidance of neurogenic niches should be considered to improve OS of HGG patients.

**Importance of Study:** Survival rates and quality of life of patients with glioma are still suboptimal, therefore improvement of radiation treatment planning and delivery is required. The SVZ and SGZ of the adult human brain are a source of brain tissue repair but may also be the source of glioma growth enhancement. By investigating the effects of radiotherapy on SVZ and SGZ, we gain insight into associations between tumor progression and survival. We included 273 adult patients with high- and low- grade glioma who received radiation treatment. We found that irradiation doses on neurogenic areas correlate with lower OS in patients with HGG. Avoidance of SVZ and SGZ should be considered to improve OS. These study results will contribute to optimization of brain tumor radiotherapy, focused on increasing OS. In order to facilitate future research into the role of the SVZ, we also provide stereotaxic standard space atlas labels for SVZ and SGZ.

## Introduction

Glioma is the most common primary brain tumor, and it consists of glial cells which normally support the functioning of nerve cells.^1,2^ Survival rates in glioma patients depend on several clinical factors, such as tumor progression and WHO grade. Patients with low-grade glioma (LGG; WHO grade I and II) have a median survival of 5.6 to 11.6 years, while patients with high-grade glioma (HGG; WHO grade III and IV) have a median survival of 14 months to 3.5 years and almost always develop recurrences.^3^ Current clinical management consists of a combination of different concurrent and consecutive treatments: tumor resection, radiotherapy (RT) and chemotherapy (CT), to reduce tumor size and inhibit progression.^2,4^ To increase survival rate and quality of life of patients with glioma, it is essential to further improve current treatment modalities and investigate the effects of modulating radiation dose to presumed tumor supporting brain regions.

Recent studies have shown that the growth of glioma might be supported by cells originating from the subventricular zone (SVZ), providing the tumor with neural stem cells (NSCs) and progenitor cells.^5–7^ Lee et al. have shown that neural tissue of HGG patients have NSCs in the SVZ that contain similar driver mutations as their matching glioma, which can migrate and result in progression of HGG.^5^ Others found that anatomical contact of the tumor with the SVZ correlates with lower survival rates in both HGG and LGG glioma, independent of other prognostic factors.^6–8^ The SVZ, which is located along the lateral wall of the ventricles, is one of the neurogenic niches in the brain, next to the subgranular zone (SGZ) in the dentate gyrus (DG) of the hippocampus (HPC).^9–11^ The main role of these niches is neurogenesis, through the generation of multipotent NSCs from the embryonic stage throughout adulthood.^9–11^ The SVZ has been shown to play a role in tissue repair and prevention of neurodegenerative diseases,^12,13^ while the SGZ generates NSCs that are involved in pathways of learning and memory.^9,14^ In contrast to NSCs from the SVZ, NSCs in the SGZ are thought unlikely to support glioma development.^15^ It has been shown that irradiation of the SGZ can result in cognitive decline in patients with central nervous system malignancies^16^, which is associated with lower overall survival (OS).^17,18^ Studies which investigated irradiation of the SVZ on the other hand, showed that HGG patients who received a high irradiation dose (>40 Gy) on the SVZ have improved OS and Progression-free Survival (PFS), compared to patients that receive a low irradiation dose on the SVZ.^19–23^ These findings suggest that for HGG, the SVZ should be targeted with high dose during RT to decrease tumor regrowth, while the SGZ might need to be spared to prevent cognitive decline. Contradictory, irradiating the SVZ and therefore damaging its repair capacity for normal brain tissue as well, may lead to opposing effects on OS as previous studies show.

The goal of this retrospective cohort study is to determine whether irradiation of the SVZ and SGZ in patients with HGG and LGG will increase survival, by comparing OS of these cohorts to irradiation doses applied to the SVZ and SGZ. By investigating the effects of radiotherapy to SVZ and SGZ in glioma patients, clues for further treatment optimization in the neuro-oncology field may be provided.

## Materials and Methods

### Patient selection and data collection

Patients diagnosed with glioma who received RT between November 2014 and July 2020 at the Department of Radiation Oncology of the UMC Utrecht were retrospectively selected. T1-weighted MR images without contrast enhancement and CT images with mapped radiation dose were collected for every patient. Gross tumor volume (GTV), clinical target volume (CTV) and planning target volume (PTV) were defined by a radiation oncologist as part of routine clinical care. Next, we collected clinical prognostic data, which includes age, sex, Karnofsky Performance Score (KPS), date of last follow-up, date of death, RT start date, survival time, RT dose, RT fractions, total intracranial volume calculated from T1 scans, (extent of) resection, (type of) CT and distance between SVZ or the HPC and GTV. The dataset also includes WHO grade and molecular markers of prognosis: isocitrate dehydrogenase (IDH)-mutation status, O6-methylguanine methyltransferase (MGMT) methylation status and 1P/19Q-codeletion status.^24–27^ Inclusion criteria for constructing the dataset were: patients diagnosed with glioma, receiving cranial RT between November 2014 and July 2020, age ≥18 years, and accessible planning and dosimetry data. Exclusion criteria were: rare cancer type or disease, re-irradiation and whole-brain radiotherapy. Patients with WHO-grade I/II and WHO-grade III/IV glioma were investigated separately as LGG and HGG cohorts, respectively. Informed consent for this retrospective study was waived by our institutional review board (#18/274).

### Image Acquisition

MR images were acquired on a 3T Philips Ingenia scanner (Philips Healthcare, Best, The Netherlands) as part of routine clinical care. T1-weighted MR images were acquired with a 3D turbo-spin echo (TSE) sequence without contrast enhancement, with the following parameters: TR = 8.1 ms, TE = 3.7 ms, flip angle = 8°, 213 continuous axial slices without gap, matrix: 207 × 289, voxel resolution 1 ×0.96×0.96mm. The planning CT scans were acquired on a Brilliance Big bore scanner (Philips Medical Systems, Best, The Netherlands), with a tube potential of 120 kVp, using a matrix size of 512 × 512 and 0.65 × 0.65 × 3.0 mm voxel size.

### Image processing and segmentation

All imaging data was processed with FSL,^28^ Statistical Parametric Mapping (SPM),^29^ Computational Anatomy Toolbox (CAT12) ^30^ and Virtual Brain Grafting (VBG).^31^ Image processing was done according to our previously published criteria.^32^ More detailed methods can be found in **Supplementary Material Appendix 1**. The SGZ masks were acquired via nonlinear registration of the Hippocampus and Subfields CoBrA atlas by Winterburn et al.^33^ using the CAT12 toolbox.^30^ Previously conducted studies acquired SVZ labels via manual delineation with varying definitions. In order to facilitate the production and reproducibility of similar research, we developed an SVZ atlas, defined in the symmetric 0.5×0.5×0.5 mm^3^ MNI-ICBM 152 space (Montreal Neurological Institute-International Consortium for Brain Mapping template created from 152 healthy brain).^34–36^ The atlas is substantiated by the neuro-anatomic development of the SVZ and divided in 4 subregions by the supervision of a neuro-pathologist. **Figure 1** shows a visual overview of the SVZ and its subregions, along with the HPC and its subregions, as well as the fornix. Details on the defining process of the SVZ atlas are available in **Supplementary Material Appendix 2**.

**Figure 1.**
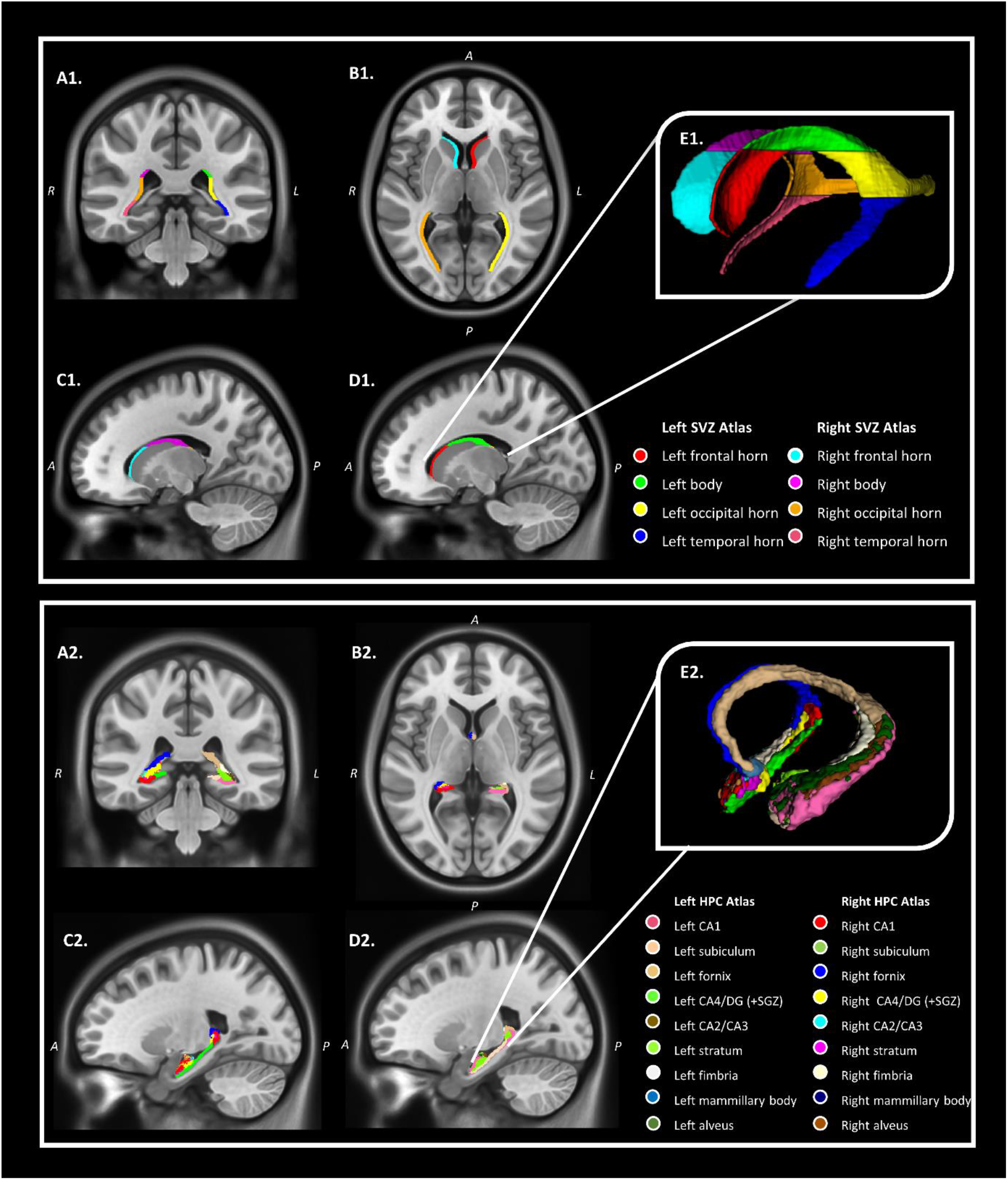
The subventricular zone (SVZ) structure (1) and hippocampus (HPC) structure (2) including the subgranular zone (SGZ) within the dentate gyrus (DG) on T1-weighted symmetrical 0.5 mm isotropic resolution template MRI images (ICBM 2009b nonlinear symmetric). **A**. Coronal view. **B**. Axial view. **C**. Sagittal view right. **D**. Sagittal view left. **E**. 3D-model of the structures, shown from the left sagittal view.

### Statistical analyses

Data analysis was performed using IBM SPSS (for Windows, version 26.0.0.1). As mentioned before, the primary outcome of this study was OS, defined as months after the first RT session until date of death or last follow-up. Patients that were alive at time of their last follow-up, are censored at the follow-up date. The relationship between the mean dose on neurogenic subregions and OS was examined using the Cox Proportional Hazards model ^37^ and the Kaplan-Meier estimator^38^. Cox Proportional Hazard models were performed with the log partial likelihood ratio test, to calculate the hazard ratios. They were executed both with and without correcting for the following prognostic factors: age, sex, KPS, total intracranial volume, mutations, surgery-extent, CT, SVZ/GTV contact and HPC/GTV contact. For the Kaplan-Meier analysis, patients were dichotomized in two groups based on the median of the mean dose for each neurogenic structure. This resulted in a high-dose group (patients that received greater than the median dose) and a low-dose group (patients that received lower than the median dose). Kaplan-Meier survival curves were compared with the log-rank test. Significance was set at *p <0*.*05* and the confidence interval (CI) was set at 95%. Moreover, Kaplan-Meier survival analysis was performed for GTV contact with the SVZ or HPC, using the same methods as Berendsen et al., Liu et al. and Chiang et al.^6–8^

## Results

### Participants

From all 338 patients that received RT between November 2014 and July 2020, a subset of 273 patients met the inclusion criteria and was selected for analysis. 32 patients were excluded due to re-irradiation and 33 patients were excluded because one or more items were inaccessible or incomplete: planning, delineation and/or dosimetry data. The remaining 273 patients were split up into HGG (n = 226) and LGG patients (n = 47). As expected, the LGG patient group did not include WHO grade I pathologies, therefore the cohort only contained WHO grade II glioma patients. The patient inclusion flowchart is shown in **Figure 2**. Baseline patient and treatment characteristics are shown in **Table 1**. In our cohort, the median OS from onset of radiotherapy of HGG patients was 12.1 months and median OS of LGG patients was 35.2 months. Median age of the HGG and LGG patients were 63 and 48 year, respectively. Prior to RT, all included patients underwent any form of surgery (biopsy, debulking, resection), and most of them received a partial resection.

**Figure 2.**
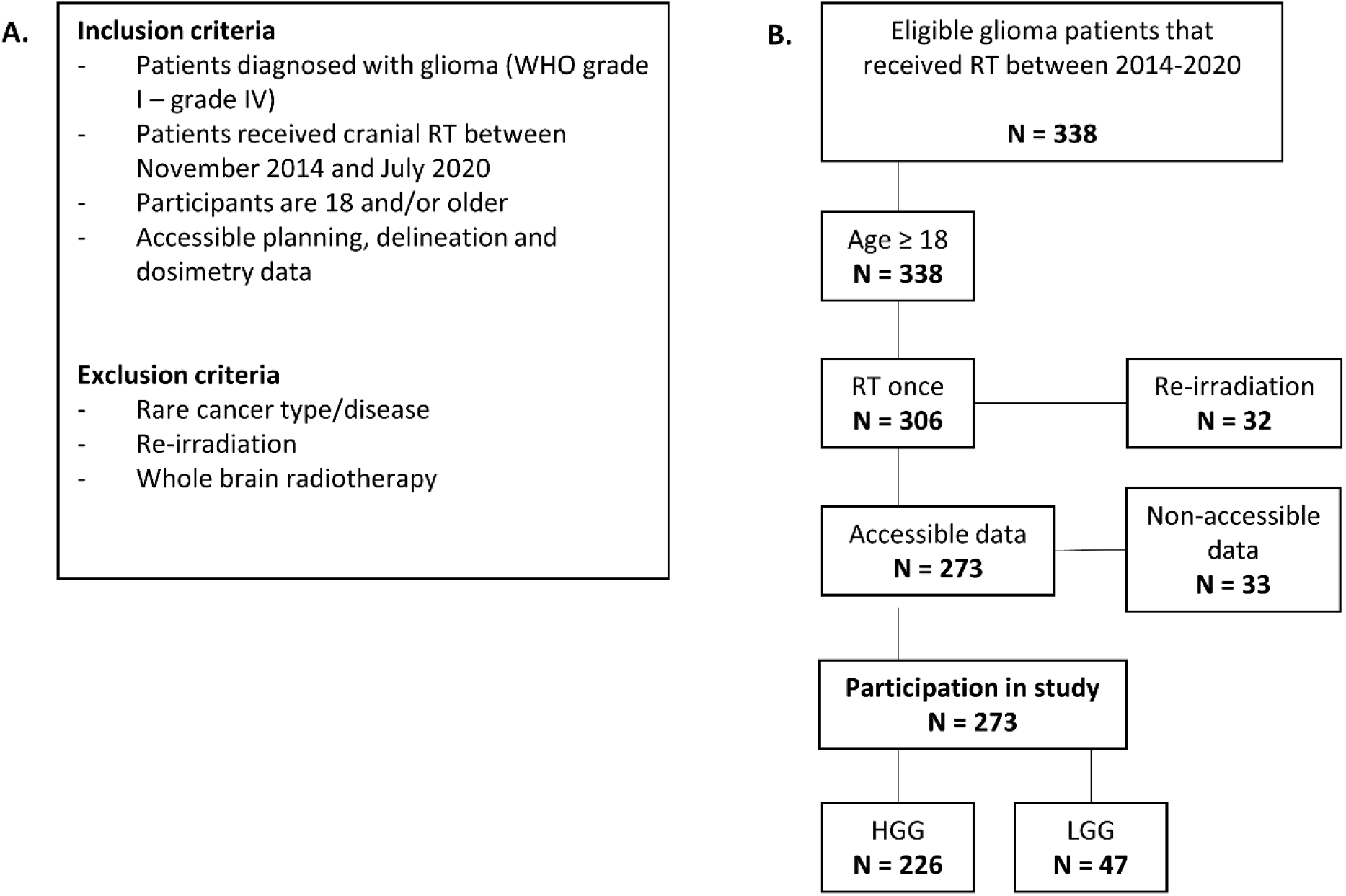
Flowchart of patient selection and overview of in- and exclusion criteria. **A**. In- and exclusion criteria for patient selection. **B**. Flowchart of glioma patient selection, divided in high-grade glioma (HGG) and low-grade glioma (LGG) patient cohorts. *RT* radiotherapy.

**Table 1.** Baseline patient and treatment characteristics of high-grade glioma (HGG) patients and low-grade glioma (LGG) patients.

| Characteristics | HGG | LGG |
| --- | --- | --- |
| Patient count (%) | 226 (100) | 47 (100) |
| Median age in years (IQR) | 63 (55-70) | 48 (37-59) |
| Sex |  |  |
| Female (%) | 79 (35) | 16 (34) |
| Male (%) | 147 (65) | 31 (66) |
| Median KPS (IQR) | 80 (70-80) | 90 (80-90) |
| Total intracranial volume in cm3 (IQR) | 1540 (1424-1632) | 1478 (1413-1666) |
| Extent of surgery |  |  |
| Biopsy (%) | 53 (23.5) | 10 (21.3) |
| Partial resection (%) | 117 (51.8) | 31 (66) |
| Complete resection (%) | 56 (24.8) | 6 (12.8) |
| Chemotherapy |  |  |
| No (%) | 42 (18.6) | 6 (12.8) |
| Yes (%) | 184 (81.4) | 41 (87.2) |
| Type of chemotherapy |  |  |
| Temozolomide (%) | 178 (96.7) | 29 (70.7) |
| Lomustine (%) | 1 (0.5) | 0 (0.0) |
| PCV (%) | 5 (2.7) | 12 (29.3) |
| GTV – SVZ contact |  |  |
| No (%) | 50 (22.1) | 12 (25.5) |
| Yes (%) | 176 (77.9) | 35 (74.5) |
| GTV – HPC contact |  |  |
| No (%) | 139 (61.5) | 22 (46.8) |
| Yes (%) | 87 (38.5) | 25 (53.2) |
| MGMT methylation |  |  |
| No (%) | 95 (42) | 3 (6.4) |
| Yes (%) | 47 (20.8) | 1 (2.1) |
| Unknown (%) | 84 (37.2) | 43 (91.5) |
| IDH mutation |  |  |
| No (%) | 208 (92) | 10 (21.3) |
| Yes (%) | 14 (6.2) | 33 (70.2) |
| Unknown (%) | 4 (1.8) | 4 (8.5) |
| 1P/19Q codeletion |  |  |
| No (%) | 105 (46.5) | 18 (38.3) |
| Yes (%) | 9 (4) | 13 (27.7) |
| Unknown (%) | 112 (49.6) | 16 (34) |

*IQR* Interquartile range, *PCV* Procarbazine Lomustine Vincristine, *SVZ* Subventricular zone, *HPC* Hippocampus, *MGMT* O6-methylguanine methyltransferase, *IDH* Isocitrate dehydrogenase.

### Survival analysis

An overview of Cox regression analyses is shown in **Table 2**, for each patient cohort separately. For HGG patients, univariate analysis resulted in a hazard ratio (HR) of 1.029 per Gy for the mean SVZ dose (*p<0*.*001*, [95% CI 1.013-1.046]) and 1.023 per Gy for the mean SGZ dose (*p<0*.*001*, [95% CI 1.013-1.033]). Multivariate analysis of HGG patient data corrected for age, sex, KPS, total intracranial volume, mutations, CT, surgery-extent and SVZ/HPC contact resulted in a HR of 1.103 (*p = 0*.*002*, [95% CI 1.037-1.173]) and 1.055 (*p<0*.*001*, [95% CI 1.027-1.083]) for the SVZ and SGZ, respectively. Dose on HPC, the occipital horn (OH) of SVZ and temporal horn (TH) of SVZ all showed significant results with an HR >1, while the frontal horn (FH) and SVZ body do not show significant results (**Table 2**).

**Table 2.** Cox regression analysis of subregion doses on overall survival (OS) in patients with high-grade glioma (HGG) and low-grade glioma (LGG). Analyses were performed univariate for irradiation dose only, and multivariate corrected for mentioned different covariates*.

| Patient diagnosis | Subregion | Univariable |  | Multivariable* |  |
| --- | --- | --- | --- | --- | --- |
|  |  | HR (95% CI) | *p-value* | HR (95% CI) | *p-value* |
| High-grade glioma | **HPC** | 1.023 (1.013-1.032) | <0.001 | 1.054 (1.026-1.083) | <0.001 |
|  | **SGZ** | 1.023 (1.012-1.033) | <0.001 | 1.055 (1.027-1.083) | <0.001 |
|  | **SVZ** | 1.029 (1.013-1.046) | <0.001 | 1.103 (1.037-1.173) | 0.002 |
|  | **FH** | 1.017 (1.004-1.030) | 0.901 | 0.996 (0.974-1.018) | 0.707 |
|  | **Body** | 1.017 (1.004-1.030) | 0.008 | 1.011 (0.968-1.056) | 0.612 |
|  | **OH** | 1.024 (1.013-1.034) | <0.001 | 1.043 (1.018-1.068) | 0.001 |
|  | **TH** | 1.024 (1.013-1.034) | <0.001 | 1.056 (1.025-1.088) | <0.001 |
|  | *Age, Sex, KPS, total intracranial volume, MGMT, IDH, 1P/19Q deletion, CT, Surgery-extent, GTV-SVZ contact or GTV-HPC contact (For the SVZ: GTV-SVZ contact, for the SGZ: GTV-SGZ contact) |  |  |  |  |
| Low-grade glioma | **HPC** | 1.033 (0.971-1.098) | 0.331 | 1.036 (0.947-1.132) | 0.441 |
|  | **SGZ** | 1.034 (0.974-1.098) | 0.273 | 1.038 (0.950-1.134) | 0.413 |
|  | **SVZ** | 1.028 (0.948-1.114) | 0.506 | 1.113 (0.949-1.305) | 0.189 |
|  | **FH** | 1.026 (0.957-1.100) | 0.470 | 1.097 (0.982-1.226) | 0.102 |
|  | **Body** | 1.002 (0.941-1.066) | 0.962 | 1.041 (0.941-1.139) | 0.385 |
|  | **OH** | 1.021 (0.963-1.082) | 0.492 | 1.014 (0.920-1.117) | 0.781 |
|  | **TH** | 1.030 (0.969-1.095) | 0.343 | 1.013 (0.935-1.097) | 0.749 |
|  | * Age, Sex, KPS, total intracranial volume, CT, Surgery-extent, GTV-SVZ contact or GTV-HPC contact (For the SVZ: GTV-SVZ contact, for the SGZ: GTV-SGZ contact) |  |  |  |  |

MGMT O6-methylguanine methyltransferase, IDH Isocitrate dehydrogenase, HPC Hippocampus, SGZ Subgranular zone, SVZ Subventricular zone, FH Frontal horn, OH Occipital horn, TH Temporal horn, HR Hazard ratio of overall survival, CI Confidence interval.

Univariate analysis for LGG patients resulted in a non-significant HR of 1.028 per Gy for mean SVZ dose (*p = 0*.*506*, [95% CI 0.948-1.114]) and 1.034 per Gy for mean SGZ dose (*p = 0*.*273*, [95% CI 0.974-1.098]). Multivariate analysis of LGG patient data could not be corrected for mutations, as patients with known mutations were still alive at time of data collection. Therefore, we corrected for age, sex, KPS, total intracranial volume, CT, surgery-extent and SVZ/HPC contact. This resulted in an HR of 1.113 (*p = 0*.*189*, [95% CI 0.949-1.305]) and 1.038 (*p = 0*.*413*, [95% CI 0.950-1.134]) for mean SVZ and SGZ doses, respectively. Both univariate and multivariate Cox regression analysis did not result in significant outcomes for any neurogenic regions in the LGG patient cohort (**Table 2**).

For Kaplan-Meier analysis, the median of the mean irradiation dose on each neurogenic subregion of the entire glioma patient cohort (HGG + LGG patients) was used as cut-off value for low- and high-dose group divisions (**Table 3**). For the SVZ, this was 30.33 Gy and for the SGZ 29.11 Gy.

**Table 3.**
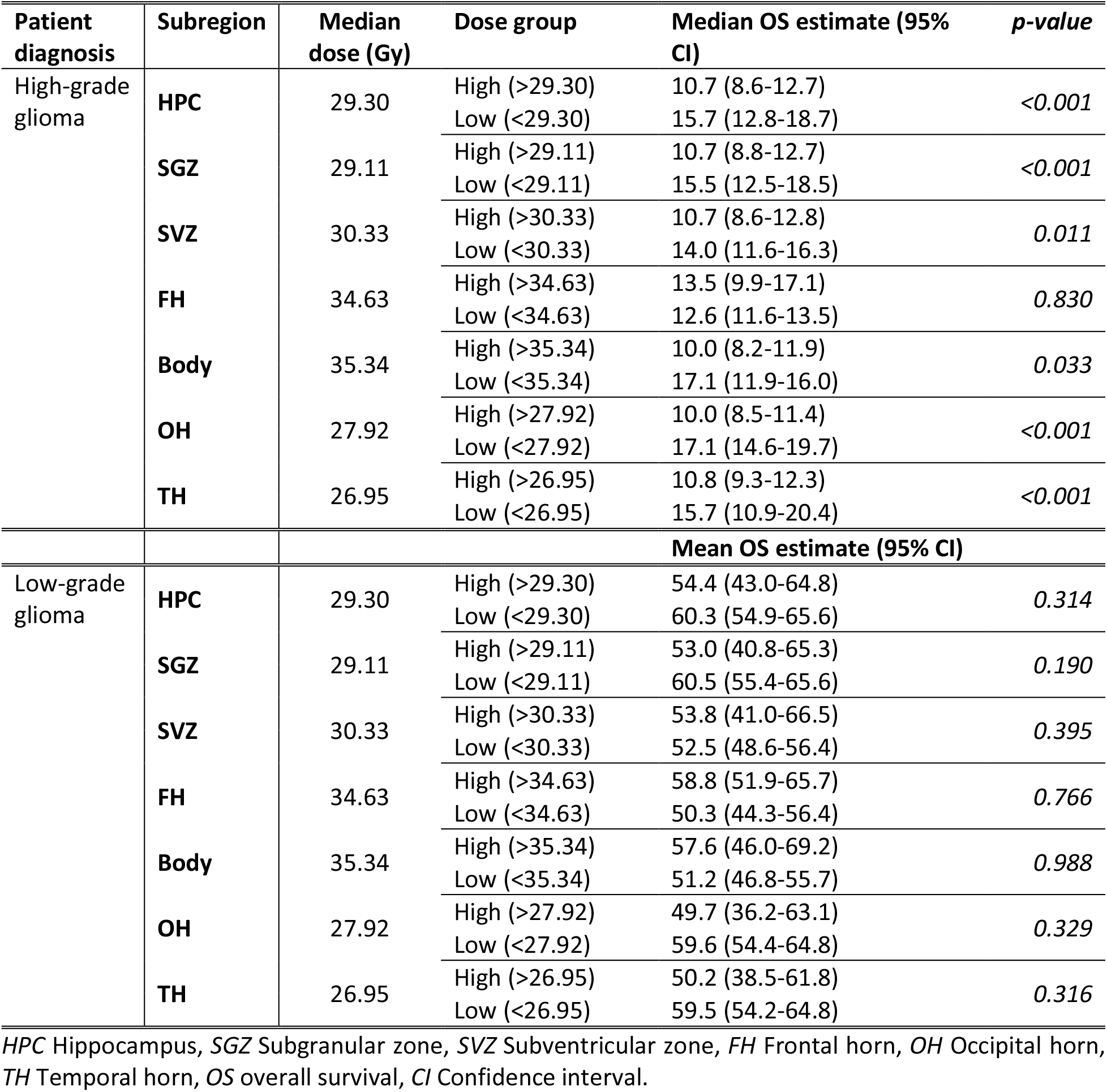
Overview of the median irradiation dose received by each neurogenic subregion and Kaplan-Meier analysis of subregion doses on overall survival (OS), with median estimates for the high-grade glioma (HGG) group, and mean estimates for low-grade glioma (LGG) patients. Estimates are given in months.

HGG patients whose SVZ received high mean SVZ dose showed a significant difference of 3.3 months shorter median OS, compared to patients who received low mean SVZ dose (10.7 months [95% CI 8.6-12.8] vs 14.0 months [95% CI 11.6-16.3] median OS, *p = 0*.*011*). In case of the SGZ, HGG patients who received high mean SGZ dose showed a significant decrease of 4.8 months in median OS, compared to patients who received low mean SGZ dose (10.7 months [95% CI 8.8-12.7] vs 15.5 months [95% CI 12.5-18.5] median OS, *p<0*.*001*).

Because the LGG cohort did not reach the proportion of 0.5 survival, median survival estimates could not be calculated and therefore only mean survival estimates are given. The corresponding Kaplan-Meier estimates and curves of SVZ and SGZ doses of both cohorts are shown in **Table 3** and **Figure 3**.

**Figure 3.**
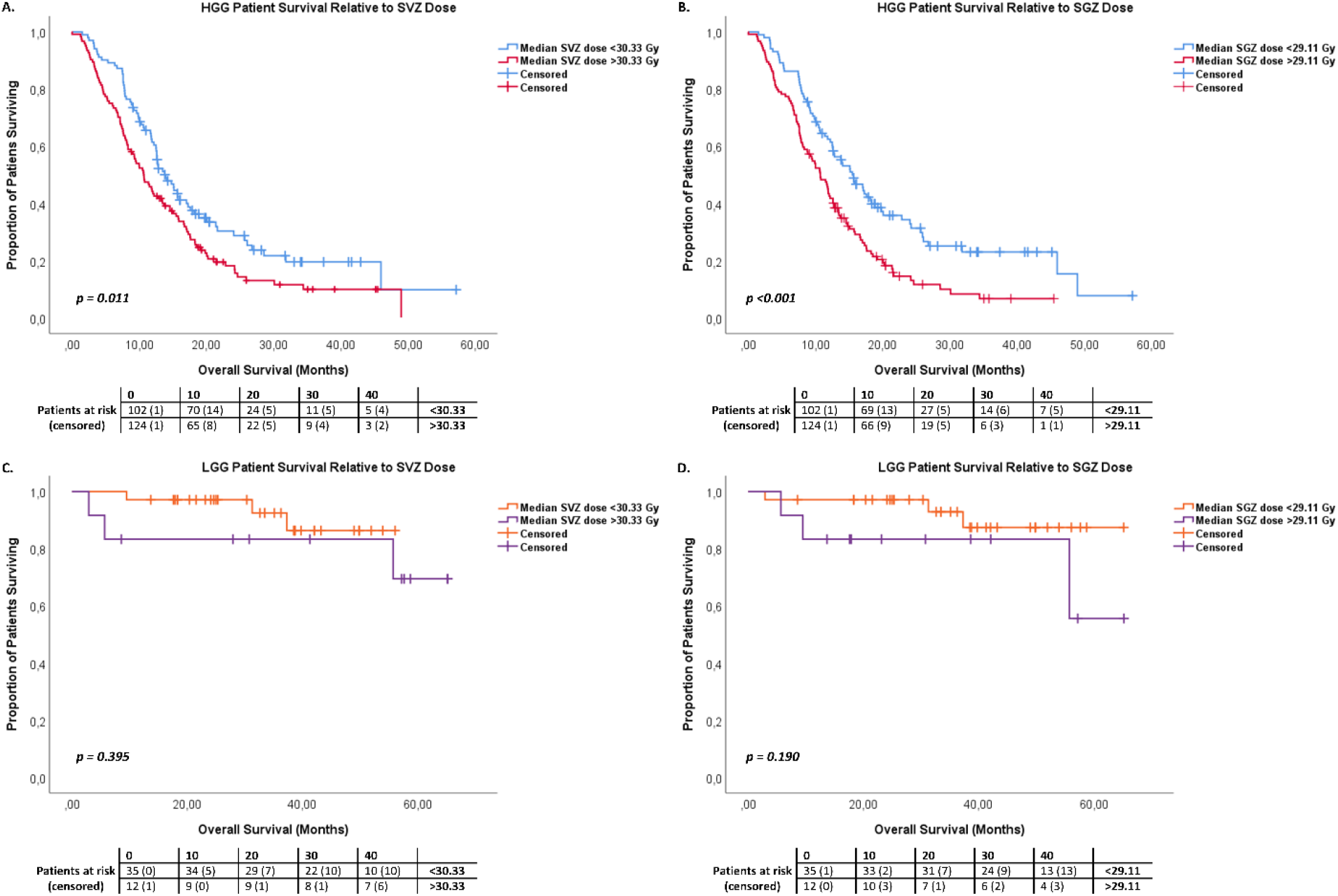
Overall survival (OS) curves for high- and low-dose groups, stratified by the median of the mean subventricular zone (SVZ) dose (A and C) and the median of the mean subgranular zone (SGZ) dose (B and D). **A**. OS curve of the proportion of high-grade glioma (HGG) patients relative to the median SVZ dose (*p = 0*.*005*, log-rank test). **B**. OS curve of the proportion of HGG patients relative to the median SGZ dose (*p<0*.*001*, log-rank test). **C**. OS curve of the proportion of low-grade glioma (LGG) patients relative to the median SVZ dose (*p = 0*.*395*, log-rank test). **D**. OS curve of the proportion of LGG patients relative to the median SGZ dose (*p = 0*.*190*, log-rank test).

For ipsilateral and contralateral SVZ and SGZ doses, LGG patients did not show any significant OS correlations. HGG patients whose ipsilateral SGZ received high mean ipsilateral SGZ dose (>45.03 Gy), showed a significant difference of 4.8 months shorter OS, compared to patients who received low mean ipsilateral SGZ dose (10.7 months [95% CI 8.1-12.5] vs 15.5 months [95% CI 12.5-18.5] median OS, *p = 0*.*002*). HGG patients whose contralateral SGZ received high mean contralateral SGZ dose (>12.54 Gy), showed a significant decrease of 5.2 months in OS, compared to patients who received low median contralateral SGZ dose (10.6 months [95% CI 9.0-12.3] vs 15.8 months [95% CI 13.1-18.6] median OS, *p = 0*.*001*). HGG patients whose ipsilateral SVZ received high mean ipsilateral SVZ dose (>39.41 Gy), showed a significant decrease of 2.3 months in OS, compared to patients who received a low mean ipsilateral SVZ dose (11.5 months [95% CI 9.7-13.4] vs 13.8 months [95% CI 11.6-16.0] median OS, *p = 0*.*035*).

HGG patients who had SVZ involvement in the GTV area had a median estimate of 4.3 months less survival compared to patients who do not have contact of the GTV with the SVZ (11.4 months [95% CI 9.7-13.1] vs 15.7 months [95% CI 13.5-17.9] median OS, *p<0*.*008*). Analyses of SVZ involvement in LGG patients did not result in significant outcomes. We also did not find correlations between HPC contact with GTV and OS, for both HGG and LGG patients.

More details and Kaplan-Meier curves of SVZ/HPC – tumor involvement, ipsilateral and contralateral dose analysis and analysis of other neurogenic subregion doses are available in **Supplementary material Appendix 3**.

## Discussion

In this retrospective cohort study, we have tested whether contact between GTV and neurogenic niches and/or irradiation dose to neurogenic niches in the adult human brain are associated with higher OS of patients with HGG and LGG. Neurogenic stem cell areas play a role in the maintenance of healthy brain tissue and repair, but may also enhance tumor growth. The hypothesis is based on previous pre-clinical and clinical research which showed that the SVZ might support the growth of LGG and HGG by providing the tumor with NSCs.^5–8,39^ However, it remains controversial, as there is a limited number of previous studies that in contrast report that irradiation of the SVZ decreases OS for patients with HGG.^40–43^ A preclinical study by Achanta et al. reported a decrease of the proliferating cell marker Ki67 in a mouse model when the SVZ was irradiated,^44^ indicating a deteriorated repair capacity. Studies from Achari et al. and Elicin et al. showed dismal effects on OS after high irradiation doses on NSC niches of patients with glioblastoma (n = 61 and n = 60 respectively),^40,43^ as did Muracciole et al. and Hallaert et al. reported for IDH-wild-type HGG patients (n = 50 and n = 137 respectively).^41,42^ Our study describes this negative correlation between SVZ irradiation and OS in HGG patients as well, involving a large number of patients (n = 226). HGG patients whose SVZ received high mean SVZ dose (>30.33 Gy), showed a significantly decrease of 3.3 months in median OS, compared to patients who received low mean SVZ dose (<30.33 Gy). Besides, multivariate Cox regression analysis showed HR’s of 1.103 and 1.055 per Gy increasing irradiation doses on the SVZ and SGZ, respectively.

On the contrary, the majority of previous studies in this field shows higher survival rates of HGG patients following irradiation of the SVZ.^19–23^ The prospective study from Malik et al. involving 45 glioblastoma patients showed improved survival when patients receive additional irradiation dose on NSC niches in the ipsilateral hemisphere.^45^ We have investigated ipsilateral and contralateral doses on neurogenic niches as well. Our retrospective outcomes show again that additional irradiation dose on both ipsilateral SVZ or SGZ is negatively correlated with OS in HGG patients, contradicting the prospective study from Malik et al. Interestingly, when considering the outcomes of the FH dose in HGG patients, neither univariate or multivariate Cox regression and Kaplan-Meier survival outcomes are significant. This may indicate that the FH is an area where the tumor is located more often than in other SVZ areas, which can interfere with the results. However, the incidence of tumor location in the patient population of this study is evenly divided between the SVZ subregions, and is not more common in the FH. This may suggest that the SVZ is not a homogeneous neurogenic region.

In contrast to the NSCs from the SVZ, previous research elucidated that NSCs in the SGZ of the HPC are not likely to sustain glioma growth, and that irradiation of the HPC may even result in cognitive decline and lower OS.^15–18^ These results are in line with our findings, where we show that HGG patients who received a high mean SGZ dose (>29.11 Gy) show a significant decrease of 4.8 months in median OS, compared to patients who received a low median SGZ dose (<29.11 Gy).

We investigated the effect of irradiation on the SVZ and SGZ in LGG patients as well. Both the univariate and the multivariate analysis of LGG patients did not result in a significant correlation between irradiation dose on neurogenic niches and OS for LGG patients. This may be due to the relatively small number of patients in our study that were diagnosed with WHO grade II.

Furthermore, we show that SVZ contact to the GTV is associated with a 4.3 months lower median OS in HGG patients compared to patients that do not have SVZ contact with the GTV, which is in line with findings by Berendsen et al.^6^ However, we are aware that this may be misleading, as the SVZ is located centrally in the brain and it is suggested that central tumor location itself is associated with worse OS.^46^ We performed similar analyses for LGG patients as well, following methods of Chiang et al.^8^ and Liu et al.^7^ but we did not find a relation between SVZ contact with GTV and survival in this patient group. The outcomes may be due to our LGG patient sample size which is less than half of the sample size used in the study of Liu et al.^7^ Another consideration is the atypical division of WHO grades in high- and low-grade, as Chiang et al.^8^ included WHO III patients as LGG patients and we included WHO III patients in the HGG cohort.

The dismal survival outcomes by high dose irradiation of neurogenic niches can be explained by the neurotoxic side-effects of irradiation outweighing the positive effects. Lower survival might be caused by the damaging of NSCs in the SVZ and SGZ, leading to decreased repair capacity and results in enhanced neurocognitive impairment, which again is associated with lower OS.^18^

The strength of our study is the high number of HGG patients included in our analyses in comparison with previous studies, which may support the soundness of our results. Also, the patients that were included received fairly homogeneous treatments, with typically surgery followed by chemoRT and adjuvant CT. Another consideration is the lack of consensus in literature on the delineation of the SVZ, due to lack of existing guidelines and/or caused by irregular manual delineations.^47^ Previous research was performed with different volumes and shapes for the SVZ, causing inconsistency in methodology among studies. In this study, we derived delineations of the SVZ from the open access SVZ atlas in standard MNI space, which now can be used in future research, enabling reproducibility. Moreover, for dichotomization in low and high RT dose groups, there is no congruence in literature about a cut-off value for irradiation dose. In addition to dichotomization in low and high RT dose groups for Kaplan-Meier analysis, we chose to include irradiation doses as continuous variables in cox-regression analyses as well, to avoid bias between specific subcategories.^48^

Nevertheless, our study shows there is room for improvements as well. First, since we could not collect clinical follow-up data other than death, PFS could not be examined. Besides, we did not assess cognitive performance of the patients, since neuropsychological evaluation was not performed routinely. Another limitation may be the effect of tumor location, because a tumor that is located near the SVZ or SGZ leads to elevated irradiation doses on the SVZ or SGZ as well. As it is suggested that central tumor location is associated with worse OS,^46^ it may interfere with the results. Therefore, we are aware that we cannot correct our outcomes for tumor location completely. Furthermore, the limited number of LGG patients in our study has probably led to insignificant results on their survival analysis. Finally, our study is of retrospective nature, which might have resulted in selection bias. To validate our results, prospective investigations are required, where irradiation and sparing of the neurogenic niches have to be compared.

In conclusion, in this retrospective study we present an SVZ atlas, SVZ delineation guidelines, and results from a large cohort of patients with HGG, that shows a statistically significant decrease in median OS with additional increased irradiation dose on the SVZ and SGZ. This suggests that, to improve OS, these neurogenic niches need to be avoided with radiotherapy in HGG patients. Modern radiotherapy planning systems and treatment delivery options are available to implement this tomorrow in most of the clinics. Nevertheless, prospective and randomized investigations in patients with glioma are required first to confirm our findings. The SVZ atlas which we defined here, is available for further use openly.

## Supporting information

SVZ Atlas

SVZ Atlas Description

## Data Availability

We share our SVZ atlas, in order to facilitate future research.

## Abbreviations

CAT: Computational Anatomy Toolbox
CI: Confidence interval
CT: Chemotherapy
CTV: Clinical target volume
DG: Dentate gyrus
FH: Frontal horn
GTV: Gross tumor volume
HGG: High-grade glioma
HPC: hippocampus
HR: Hazard ratio
IDH: Isocitrate dehydrogenase
IQR: Interquartile range
KPS: Karnofsky Performance Score
LGG: Low-grade glioma
MGMT: O6-Methylguanine methyltransferase
NSC: Neural stem cell
OH: Occipital horn
OS: Overall survival
PFS: Progression-free survival
PTV: planning target volume
RT: radiotherapy
SGZ: Subgranular zone
SPM: Statistical Parametric Mapping
SVZ: Subventricular zone
TH: Temporal horn
TSE: Turbo Spin Echo
VBG: Virtual Brain Grafting

## Acknowledgements

We would like to thank Dr. Angelika Mühlebner (Department of Pathology, UMC Utrecht) for her contributions to the SVZ atlas.

## References

1 Vescovi, Angelo L, Galli, Rossella and Reynolds, Brent A (2006) “Brain tumour stem cells.” Nature Reviews Cancer, 6(6), pp. 425–436.

2 Reijneveld, J C (2010) “Neuro-oncologie,” in Het Neurologie Formularium, Springer, pp. 210–217.

3 Sizoo, E M, Reijneveld, J C, Lagerwaard, F J, Buter, J, et al. (2010) “Beloop en beleid bij vermoeden van een laaggradig glioom.” Nederlands Tijdschrift voor Geneeskunde, 154.

4 Stupp, Roger, Mason, Warren P, van den Bent, Martin J, Weller, Michael, et al. (2005) “Radiotherapy plus concomitant and adjuvant temozolomide for glioblastoma.” New England journal of medicine, 352(10), pp. 987–996.

5 Lee, Joo Ho, Lee, Jeong Eun, Kahng, Jee Ye, Kim, Se Hoon, et al. (2018) “Human glioblastoma arises from subventricular zone cells with low-level driver mutations.” Nature, 560(7717), pp. 243–247.

6 Berendsen, Sharon, van Bodegraven, Emma, Seute, Tatjana, Spliet, Wim G M, et al. (2019) “Adverse prognosis of glioblastoma contacting the subventricular zone: Biological correlates.” PloS one, 14(10), p. e0222717.

7 Liu, Shuai, Wang, Yinyan, Fan, Xing, Ma, Jun, et al. (2016) “Anatomical Involvement of the Subventricular Zone Predicts Poor Survival Outcome in Low-Grade Astrocytomas.” PLOS ONE, 11(4), p. e0154539.

8 Chiang, Gloria C, Pisapia, David J, Liechty, Benjamin, Magge, Rajiv, et al. (2020) “The Prognostic Value of MRI Subventricular Zone Involvement and Tumor Genetics in Lower Grade Gliomas.” Journal of Neuroimaging, 30(6), pp. 901–909.

9 Zhao, Chunmei, Deng, Wei and Gage, Fred H (2008) “Mechanisms and functional implications of adult neurogenesis.” Cell, 132(4), pp. 645–660.

10 Katsimpardi, Lida and Lledo, Pierre-Marie (2018) “Regulation of neurogenesis in the adult and aging brain.” Current opinion in neurobiology, 53, pp. 131–138.

11 Bond, Allison M, Ming, Guo-li and Song, Hongjun (2015) “Adult mammalian neural stem cells and neurogenesis: five decades later.” Cell stem cell, 17(4), pp. 385–395.

12 Richardson, R Mark, Sun, Dong and Bullock, M Ross (2007) “Neurogenesis after traumatic brain injury.” Neurosurgery Clinics, 18(1), pp. 169–181.

13 Curtis, Maurice A, Faull, Richard L M and Eriksson, Peter S (2007) “The effect of neurodegenerative diseases on the subventricular zone.” Nature Reviews Neuroscience, 8(9), pp. 712–723.

14 Deng, Wei, Aimone, James B and Gage, Fred H (2010) “New neurons and new memories: how does adult hippocampal neurogenesis affect learning and memory?” Nature reviews neuroscience, 11(5), pp. 339–350.

15 Mistry, Akshitkumar M, Dewan, Michael C, White-Dzuro, Gabrielle A, Brinson, Philip R, et al. (2017) “Decreased survival in glioblastomas is specific to contact with the ventricular-subventricular zone, not subgranular zone or corpus callosum.” Journal of neuro-oncology, 132(2), pp. 341–349.

16 Gondi, Vinai, Tomé, Wolfgang A and Mehta, Minesh P (2010) “Why avoid the hippocampus? A comprehensive review.” Radiotherapy and Oncology, 97, pp. 370–376.

17 Taphoorn, Martin J.B. and Klein, Martin (2004) “Cognitive deficits in adult patients with brain tumours.” Lancet Neurology, 3(3), pp. 159–168.

18 Klein, M, Postma, T J, Taphoorn, M J B, Aaronson, N K, et al. (2003) “The prognostic value of cognitive functioning in the survival of patients with high-grade glioma.” Neurology, 61(12), pp. 1796–1798.

19 Darázs, Barbara, Ruskó, László, Végváry, Zoltán, Ferenczi, Lehel, et al. (2019) “Subventricular zone volumetric and dosimetric changes during postoperative brain tumor irradiation and its impact on overall survival.” Physica Medica, 68, pp. 35–40.

20 Chen, Linda, Guerrero-Cazares, Hugo, Ye, Xiaobu, Ford, Eric, et al. (2013) “Increased subventricular zone radiation dose correlates with survival in glioblastoma patients after gross total resection.” International Journal of Radiation Oncology* Biology* Physics, 86(4), pp. 616–622.

21 Lee, Percy, Eppinga, Wietse, Lagerwaard, Frank, Cloughesy, Timothy, et al. (2013) “Evaluation of high ipsilateral subventricular zone radiation therapy dose in glioblastoma: a pooled analysis.” International Journal of Radiation Oncology* Biology* Physics, 86(4), pp. 609–615.

22 Evers, Patrick, Lee, Percy P, DeMarco, John, Agazaryan, Nzhde, et al. (2010) “Irradiation of the potential cancer stem cell niches in the adult brain improves progression-free survival of patients with malignant glioma.” BMC Cancer 2010 10:1, 10(1), pp. 1–7.

23 Gupta, Tejpal, Nair, Vimoj, Paul, Siji Nojin, Kannan, Sadhana, et al. (2012) “Can irradiation of potential cancer stem-cell niche in the subventricular zone influence survival in patients with newly diagnosed glioblastoma?” Journal of Neuro-Oncology 2012 109:1, 109(1), pp. 195–203.

24 Prognostik, Düflük Evreli Gliomlarda (2008) “Efficacy of prognostic factors on survival in patients with low grade glioma.” Turkish neurosurgery, 18(4), pp. 336–344.

25 Thakkar, Jigisha P, Dolecek, Therese A, Horbinski, Craig, Ostrom, Quinn T, et al. (2014) “Epidemiologic and molecular prognostic review of glioblastoma.” Cancer Epidemiology and Prevention Biomarkers, 23(10), pp. 1985–1996.

26 Zhao, Ye-Yu, Chen, Si-Hai, Hao, Zheng, Zhu, Hua-Xin, et al. (2019) “A nomogram for predicting individual prognosis of patients with low-grade glioma.” World neurosurgery, 130, pp. e605–e612.

27 Ducray, François Idbaih, Ahmed, Wang, Xiao-Wei, Cheneau Caroline, et al. (2011) “Predictive and prognostic factors for gliomas.” Expert review of anticancer therapy, 11(5), pp. 781–789.

28 Jenkinson, Mark, Beckmann, Christian F., Behrens, Timothy E.J., Woolrich, Mark W. and Smith, Stephen M. (2012) “FSL.” NeuroImage, 62(2), pp. 782–790.

29 Penny, William D, Friston, Karl J, Ashburner, John T, Kiebel, Stefan J and Nichols, Thomas E (2011) Statistical parametric mapping: the analysis of functional brain images, Elsevier.

30 Gaser, C, Hbm, R Dahnke - and 2016, undefined (n.d.) “CAT-a computational anatomy toolbox for the analysis of structural MRI data.” neuro.uni-jena.de.

31 Radwan, Ahmed M., Emsell, Louise, Blommaert, Jeroen, Zhylka, Andrey, et al. (2021) “Virtual brain grafting: Enabling whole brain parcellation in the presence of large lesions.” NeuroImage, 229, p. 117731.

32 Nagtegaal, Steven H.J., David, Szabolcs, van der Boog, Arthur T.J., Leemans, Alexander and Verhoeff, Joost J.C. (2019) “Changes in cortical thickness and volume after cranial radiation treatment: A systematic review.” Radiotherapy and Oncology, 135, pp. 33–42.

33 Winterburn, Julie L., Pruessner, Jens C., Chavez, Sofia, Schira, Mark M., et al. (2013) “A novel in vivo atlas of human hippocampal subfields using high-resolution 3T magnetic resonance imaging.” NeuroImage, 74, pp. 254–265.

34 Fonov, Vladimir, Evans, Alan C., Botteron, Kelly, Almli, C. Robert, et al. (2011) “Unbiased average age-appropriate atlases for pediatric studies.” NeuroImage, 54(1), pp. 313–327.

35 Fonov, VS, Evans, AC, McKinstry, RC, Almli, CR and Collins, DL (2009) “Unbiased nonlinear average age-appropriate brain templates from birth to adulthood.” NeuroImage, 47, p. S102.

36 Atlas, ICBM (2001) “McConnell Brain Imaging Centre.” Montréal Neurological Institute, McGill University, Montréal, Canada.

37 Cox, David R (1972) “Regression models and life-tables.” Journal of the Royal Statistical Society: Series B (Methodological), 34(2), pp. 187–202.

38 Kaplan, Edward L and Meier, Paul (1958) “Nonparametric estimation from incomplete observations.” Journal of the American statistical association, 53(282), pp. 457–481.

39 Marsh, James C, Wendt, Julie A, Herskovic, Arnold M, Diaz, Aidnag, et al. (2012) “High-grade glioma relationship to the neural stem cell compartment: a retrospective review of 104 cases.” International Journal of Radiation Oncology* Biology* Physics, 82(2), pp. e159–e165.

40 Achari, R., Arunsingh, M., Badgami, R. K., Saha, A., et al. (2017) “High-dose Neural Stem Cell Radiation May Not Improve Survival in Glioblastoma.” Clinical Oncology, 29(6), pp. 335–343.

41 Muracciole, Xavier, El-Amine, Wassim, Tabouret, Emmeline, Boucekine, Mohamed, et al. (2018) “Negative survival impact of high radiation doses to neural stem cells niches in an IDH-wild-type glioblastoma population.” Frontiers in oncology, 8, p. 426.

42 Hallaert, Giorgio, Pinson, Harry, van den Broecke, Caroline, Sweldens, Caroline, et al. (2021) “Survival impact of incidental subventricular zone irradiation in IDH-wildtype glioblastoma.” Acta Oncologica, 60(5), pp. 613–619.

43 Elicin, Olgun, Inac, Ebrar, Uzel, Esengul Kocak, Karacam, Songul and Uzel, Omer Erol (2014) “Relationship between survival and increased radiation dose to subventricular zone in glioblastoma is controversial.” Journal of neuro-oncology, 118(2), pp. 413–419.

44 Achanta, Pragathi, Capilla-Gonzalez, Vivian, Purger, David, Reyes, Juvenal, et al. (2012) “Subventricular zone localized irradiation affects the generation of proliferating neural precursor cells and the migration of neuroblasts.” Stem Cells, 30(11), pp. 2548–2560.

45 Malik, M., Akram, K.S., Joseph, D., Valiyaveettil, D. and Ahmed, S.F. (2015) “Prospective Study of Irradiation of Potential Stem Cell Niches in Glioblastoma.” International Journal of Radiation Oncology*Biology*Physics, 93(3), p. S111.

46 Fyllingen, Even Hovig, Bø, Lars Eirik, Reinertsen, Ingerid, Store Jakola, Asgeir, et al. (2021) “Survival of glioblastoma in relation to tumor location: a statistical tumor atlas of a population- based cohort.” Acta Neurochirurgica, 163, pp. 1895–1905.

47 Nourallah, B., Digpal, R., Jena, R. and Watts, C. (2017) “Irradiating the Subventricular Zone in Glioblastoma Patients: Is there a Case for a Clinical Trial?” Clinical Oncology, 29(1), pp. 26–33.

48 Royston, Patrick, Altman, Douglas G and Sauerbrei, Willi (2006) “Dichotomizing continuous predictors in multiple regression: a bad idea.” STATISTICS IN MEDICINE Statist. Med, 25(1), pp. 127–141.

