## Supplementary Material for "Irradiation of the Subventricular Zone and Subgranular Zone: an Atlas-based analysis on Overall Survival in High- and Low-Grade Glioma Patients"

**Abbreviations**

CSF = Cerebrospinal fluid; FOV = Field-of-view; GM = Gray matter; MRF = Markov Random Field; SANLM = Spatial-adaptive nonlocal means; WM = White matter

**Appendix 1, Methods – Image Processing**

The field-of-view (FOV) of the CT image was reduced to include only skull and upper parts of the neck to aid the performance of further image processing. The FOV-reduced CT and the associated dose and the GTV, CTV and PTV maps were registered rigidly (using 6 degrees of freedom) to the T1 MR images using FSL tool FLIRT.[1–3] This step resulted in the CT image along with all the RT data and the MRIs in the same coordinate system with identical voxel sizes. Tissue segmentation and atlas registration can be challenging in the surgically and radiation treated brains, because it is common to find residual damage (e.g. edema, surgical scarring, tumor bed, etc.). In order to ensure the successful SVZ and SGZ atlas registration, that is not affected by the aforementioned damaged brain tissue, we applied virtual brain grafting (VBG)[4] on all patients' brains to replace the damaged areas with normal-appearing tissue. VBG is an open-source, automated process in which a predefined 'lesion area' is filled with anatomically meaningful tissue, while ensuring that the filling tissue's intensity and noise profile is similar to its surroundings. We used the T1-coregistered GTVs to perform the tissue replacement, and therefore manual definition of the damaged area was not necessary. In case of 4 patients, the extent of brain damage was larger than the GTV. Therefore, we used the CTV for lesion-filling in those cases. The application of the VBG process is a critical image processing step, because none of the commonly used brain image processing softwares - FSL,[5] Freesurfer,[6] SPM,[7] and CAT[8] - can handle other tissue that is included in their anatomical models: white matter (WM), gray matter (GM) and cerebrospinal fluid (CSF). Therefore, the blind application of such tools can result in incomplete or failed segmentation/registration. Next, the VGB-enhanced T1 images were processed with CAT12's standard, single image segmentation pipeline (version 12.7 r1742), using the default options. The package includes bias-field inhomogeneity correction, spatial-adaptive non-local means (SANLM) denoising,[9] tissue segmentation to GM, WM and CSF[10] and spatially normalization using the DARTEL algorithm.[11] The segmentations were further finetuned by accounting for partial volume effects[12] by using a hidden Markov Random Field (MRF) model.[12] Whole brain TIV was calculated, along with the volumes of SVZ and HPC regions and subregions. In case of overlap of the GTV and any of the (sub)regions, we only counted the non-overlapping parts as region volume. Also, when GTV overlapped with any of the regions, it was recorded as 'tumor involvement'. Labels from the CoBrA atlas were provided by CAT12 natively, while our currently defined SVZ atlas was used via the 'own atlas maps' option to acquire the SVZ labels in the native T1 images. After the complete atlas registration, the atlas labels which cover the lesion-filled area were replaced with a 'lesion' tag and the filled areas were not used in any subsequent analysis. Mean dose for every region and subregion was calculated from the T1-coregistered dose maps and the delineations provided by CAT12. **Supplementary Figure 1** gives an overview of the image processing pipeline.

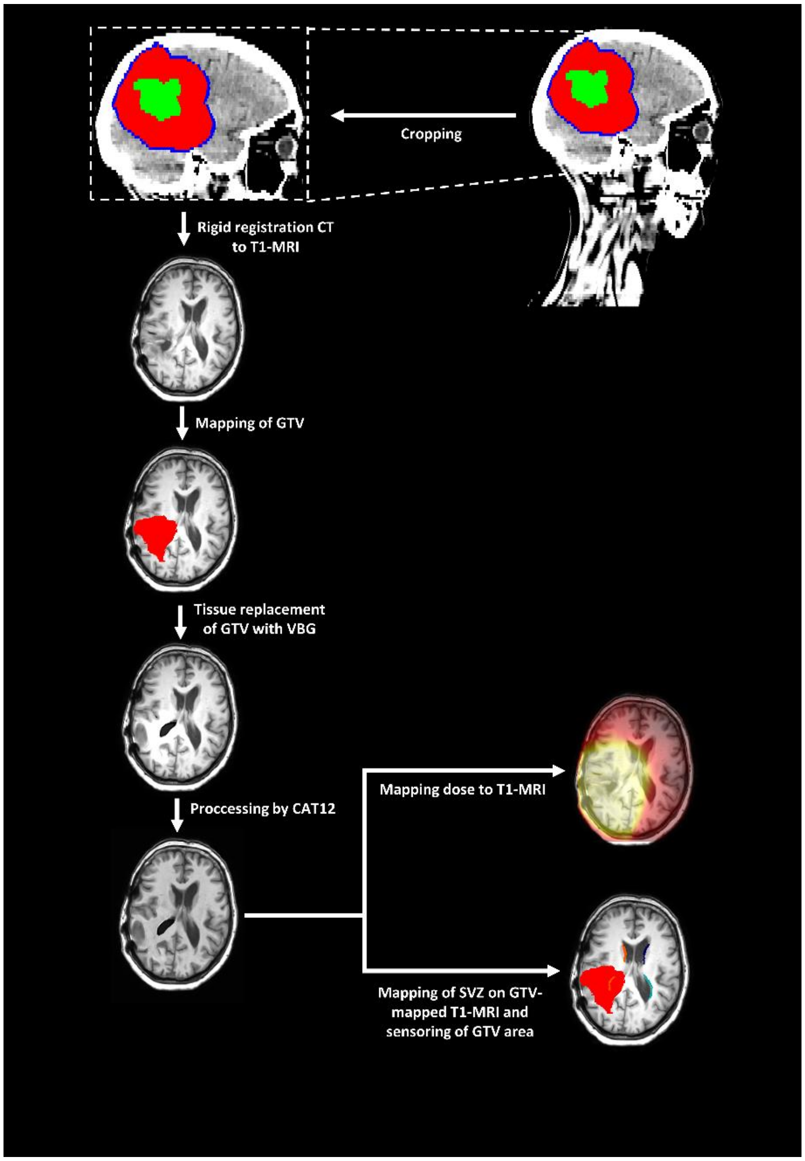


**Supplementary Figure 1.** Image processing pipeline of mapping the subventricular zone (SVZ) and subgranular zone (SGZ), censoring of gross tumor volume (GTV) area, and mapping radiation dose to Virtual Brain Grafting (VBG)-enhanced T1 MRI.

**Appendix 2, Methods - Manual Segmentation of the Subventricular Zone**

To calculate the mean dose from the SVZ and SGZ areas in an automated and reproducible fashion, accurate labels are required. To this end, we used the SGZ labels from the Hippocampus and Subfields CoBrA atlas by Winterburn et al.[13] (**Manuscript Figure 1**). To the best of our knowledge, the SVZ labels are not described in any standardized brain atlases. Therefore, we manually created the SVZ segmentation, based-on the T1- and T2- weighted, symmetrical, 0.5x 0.5x0.5 $mm^3$ resolution brain templates, available in MNI space. The atlases are freely available from the MNI website: https://www.bic.mni.mcgill.ca/ServicesAtlases/ICBM152NLin2009 (ICBM 2009b Nonlinear Symmetric 0.5×0.5×0.5mm template). We used the following software for delineation: ITKsnap 3.6.0, obtained from itksnap.org, and MRIcron, obtained from www.nitrc.org/projects/mricron.

The SVZ is a very thin area located between the lateral ventricles and the caudate nucleus. Based on the neuroanatomy of the SVZ, previous studies[14–18], and expert opinion, the thickness of the SVZ in this segmentation has been established at a margin of 3 mm along the lateral walls of the ventricles. The segmentation followed the thin space between the ventricles and caudate nucleus, and was based on the embryonal development of the SVZ anatomy.[19] Initial labelling was performed in axial orientation and was adjusted in sagittal and coronal dimensions.

To be as concise as possible concerning the tumor site, both left and right SVZ were divided into four different subregions, according to A.L. Rhoton.[20] The subregions are divided into the frontal horn (FH), body, occipital horn (OH) and temporal horn (TH) of the SVZ. This anatomical division has led to the final SVZ atlas as shown in **Manuscript Figure 1**. Anatomically, the interventricular foramen indicates the border between the frontal horn and body subregion (**Supplementary Figure 2**), as the frontal horn is located anterior to the interventricular foramen in the frontal lobe, and the body is located posterior in the parietal lobe, close to the corpus callosum and fornix.[20] The occipital horn subregion expands into the occipital lobe, and outlines the bulb of the corpus callosum.[20] The temporal lobe contains the temporal horn subregion, which lies close to the HPC.[20] **Supplementary Figure 3** shows examples of patient scans and their SVZ segmentations that were acquired in our study. Label colors are different from our original SVZ atlas that we show in our paper.

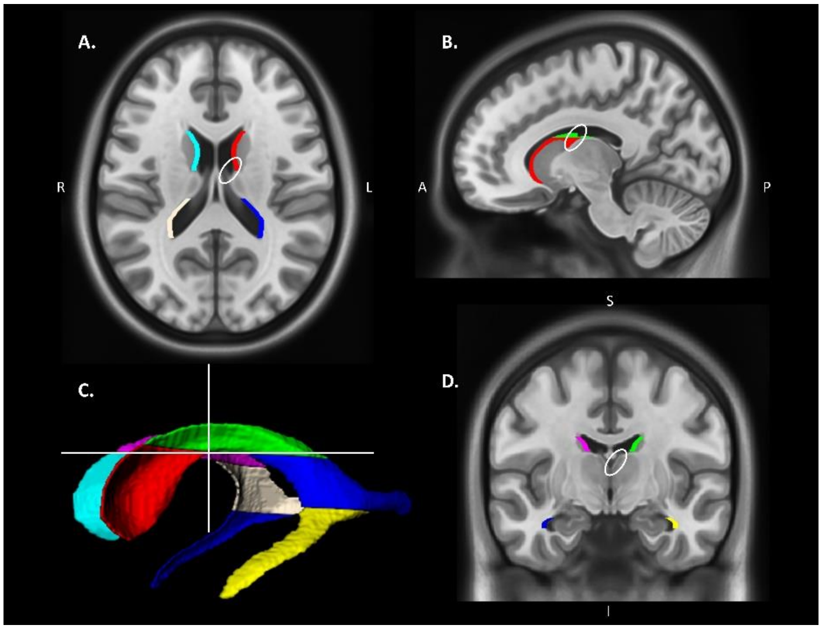


**Supplementary Figure 2.** The intraventricular foramen indicates the border between the frontal horn (red and light blue labelled) and body (green and purple labelled) of the subventricular zone (SVZ), encircled in white in axial (**A**), sagittal (**B**) and coronal (**D**) view. The intersection shown in the 3D-model of the SVZ (**C**), also shows where the intraventricular septum is located.

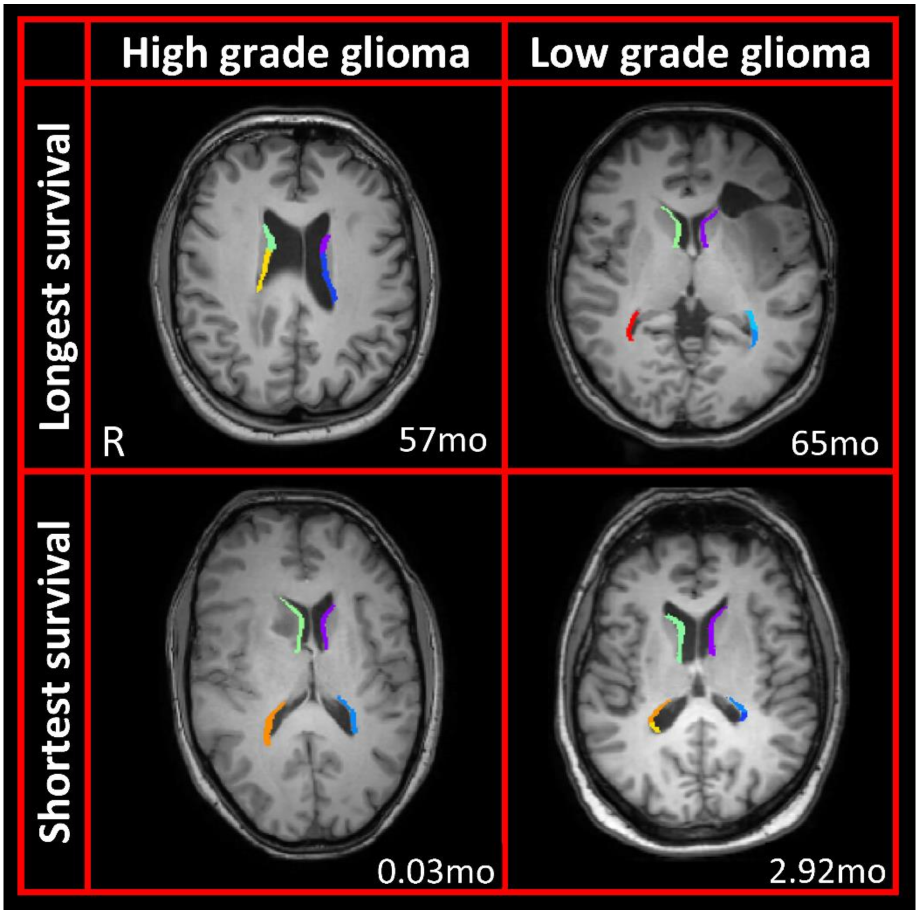


**Supplementary Figure 3.** Example T1 MRI scans and SVZ segmentations in the presence of low-grade glioma (LGG) or high-grade glioma (HGG) for the shortest and longest surviving patients. Survival is shown in months (mo).

**Appendix 3, Results**

**Supplementary Table 1** shows the correction process of Cox regression analyses. Univariate analysis and the most important multivariate analysis are shown in **Manuscript Table 2**. **Supplementary Table 2** and **Supplementary Figure 4** show results of SVZ/HPC and GTV contact.

**Manuscript Table 3** of the main text shows the Kaplan-Meier overall survival (OS) estimates of all the different neurogenic subregions in both HGG and LGG patient cohorts. **Supplementary Figure 5** (HGG) and **Supplementary Figure 6** (LGG) show the corresponding Kaplan-Meier curves for the HPC and FH, body, OH and TH of the SVZ.

We provide additional information about ipsilateral and contralateral irradiation doses on the SVZ and SGZ. An overview of Cox regression analysis of these doses for both patient groups is shown in **Supplementary Table 3**. For HGG patients, all univariate as well as multivariate results are significant and have an HR>1, which indicates that dose on both ipsilateral or contralateral regions affect survival outcomes negatively. For LGG patients, neither univariate or multivariate Cox regression analyses resulted in significant outcomes.

Kaplan-Meier analysis was performed as well, in which the median dose values are used to stratify the doses into a high and a low dose group. Results of these analyses are available in **Supplementary Table 4**. For LGG patients, ipsi- and contralateral SVZ doses do not correlate with OS. For HGG patients, irradiation of the ipsilateral and contralateral SGZ, and the ipsilateral SVZ, shows a negative association. The corresponding survival curves are shown in **Supplementary Figure 7** and **Supplementary Figure 8**.

**Supplementary Table 1.** Cox regression analysis of subregion doses on overall survival (OS) in patients with high-grade glioma (HGG) and low-grade glioma (LGG). Analyses were performed univariate for irradiation dose only, and multivariate corrected for mentioned different covariates.

| Patient diagnosis | Corrected for covariates | | | HPC | SGZ | SVZ | FH | Body | OH | TH |
|---|---|---|---|---|---|---|---|---|---|---|
| High-grade glioma | Age, Sex, KPS, total intracranial volume | | *p-value* | *<0.001* | *<0.001* | *<0.001* | *0.934* | *0.005* | *<0.001* | *<0.001* |
| | | | HR | 1.026 | 1.026 | 1.036 | 1.000 | 1.019 | 1.026 | 1.027 |
| | | 95.0% CI for HR | Lower | 1.016 | 1.016 | 1.019 | 0.992 | 1.006 | 1.015 | 1.016 |
| | | | Upper | 1.036 | 1.037 | 1.055 | 1.009 | 1.033 | 1.038 | 1.039 |
| | Age, Sex, KPS, total intracranial volume, MGMT, IDH, 1P/19Q deletion, Chemotherapy, Surgery-extent | | *p-value* | *0.001* | *<0.001* | *0.002* | *0.869* | *0.385* | *0.001* | *<0.001* |
| | | | HR | 1.048 | 1.049 | 1.083 | 1.002 | 1.017 | 1.043 | 1.057 |
| | | 95.0% CI for HR | Lower | 1.021 | 1.021 | 1.029 | 0.983 | 0.979 | 1.018 | 1.027 |
| | | | Upper | 1.077 | 1.076 | 1.141 | 1.021 | 1.058 | 1.069 | 1.087 |
| Low-grade glioma | Age, Sex, KPS, total intracranial volume | | *p-value* | *0.689* | *0.658* | *0.392* | *0.137* | *0.805* | *0.702* | *0.708* |
| | | | HR | 1.012 | 1.013 | 1.042 | 1.063 | 1.008 | 1.014 | 1.012 |
| | | 95.0% CI for HR | Lower | 0.954 | 0.956 | 0.948 | 0.981 | 0.945 | 0.945 | 0.950 |
| | | | Upper | 1.074 | 1.074 | 1.145 | 1.151 | 1.075 | 1.088 | 1.079 |
| | Age, Sex, KPS, total intracranial volume, Chemotherapy, Surgery-extent | | *p-value* | *0.755* | *0.709* | *0.193* | *0.054* | *0.365* | *0.533* | *0.722* |
| | | | HR | 1.012 | 1.014 | 1.088 | 1.110 | 1.040 | 1.028 | 1.014 |
| | | 95.0% CI for HR | Lower | 0.938 | 0.942 | 0.958 | 0.998 | 0.955 | 0.943 | 0.940 |
| | | | Upper | 1.092 | 1.093 | 1.236 | 1.236 | 1.132 | 1.120 | 1.093 |

*MGMT* O6-methylguanine methyltransferase, *IDH* Isocitrate dehydrogenase, *HPC* Hippocampus, *SGZ* Subgranular zone, *SVZ* Subventricular zone, *FH* Frontal horn, *OH* Occipital horn, *TH* Temporal horn, *HR* Hazard ratio of overall survival, *CI* Confidence interval.

**Supplementary Table 2.** Overview of Kaplan-Meier analysis of contact between the tumor and neurogenic region on overall survival (OS), with median and mean estimates for the high-grade glioma (HGG) group, and mean estimates for the low-grade glioma (LGG) group. Estimates are given in months.

| **Patient diagnosis** | | | **SVZ involvement** | | **HPC involvement** | |
|---|---|---|---|---|---|---|
| | **Contact between tumor and brain region** | | **Yes** | **No** | **Yes** | **No** |
| HGG patients | Median OS estimate (months) | | 11.4 | 15.7 | 12.2 | 12.8 |
| | 95,0% CI | Lower | 9.7 | 13.5 | 10.9 | 10.4 |
| | | Upper | 13.1 | 17.9 | 13.5 | 15.2 |
| | Mean OS estimate (months) | | 16.0 | 22.1 | 18.8 | 18.1 |
| | 95.0% CI | Lower | 13.7 | 17.5 | 14.4 | 15.1 |
| | | Upper | 18.3 | 26.7 | 21.3 | 21.2 |
| | *p-value* | | *0.008* | | *0.957* | |
| LGG patients | Mean OS estimate (months) | | 57.4 | 62.9 | 54.1 | 62.9 |
| | 95.0% CI | Lower | 51.2 | 58.9 | 45.5 | 58.9 |
| | | Upper | 63.6 | 66.9 | 62.8 | 66.9 |
| | *p-value* | | *0.678* | | *0.113* | |

*SVZ* Subventricular zone, HPC Hippocampus, *OS* Overall survival, *CI* Confidence interval.

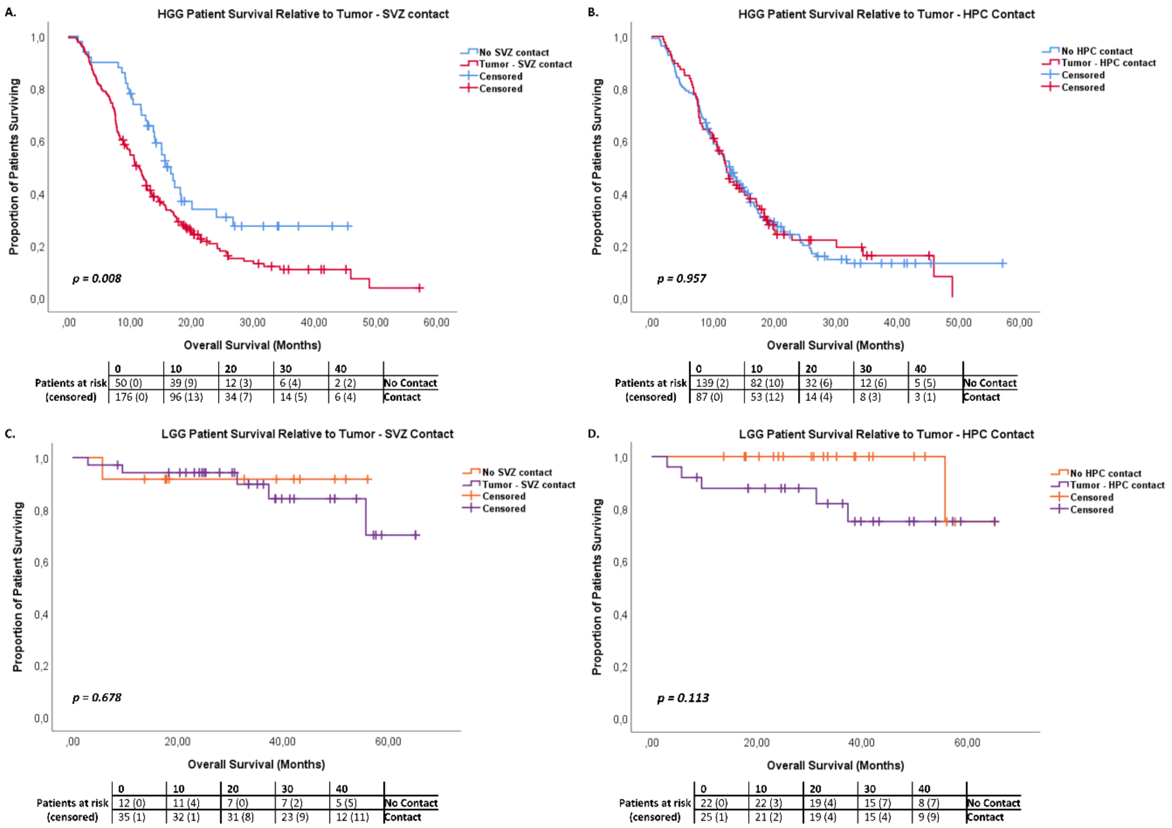


**Supplementary Figure 4.** Patient survival curves of tumor contact with subventricular zone (SVZ) and hippocampus (HPC) in low-grade glioma (LGG) and high-grade glioma (HGG) patients, that illustrate the difference in overall survival (OS) between tumor contact with the neurogenic zone or not. **A1.** HGG patient survival curve relative to tumor contact with the SVZ (*p = 0.008*). **B1.** HGG patient survival curve relative to tumor contact with the HPC (*p = 0.957*). **A2.** LGG patient survival curve relative to tumor contact with the SVZ (*p = 0.678*). **B2.** LGG patient survival curve relative to tumor contact with the HPC (*p = 0.113*).

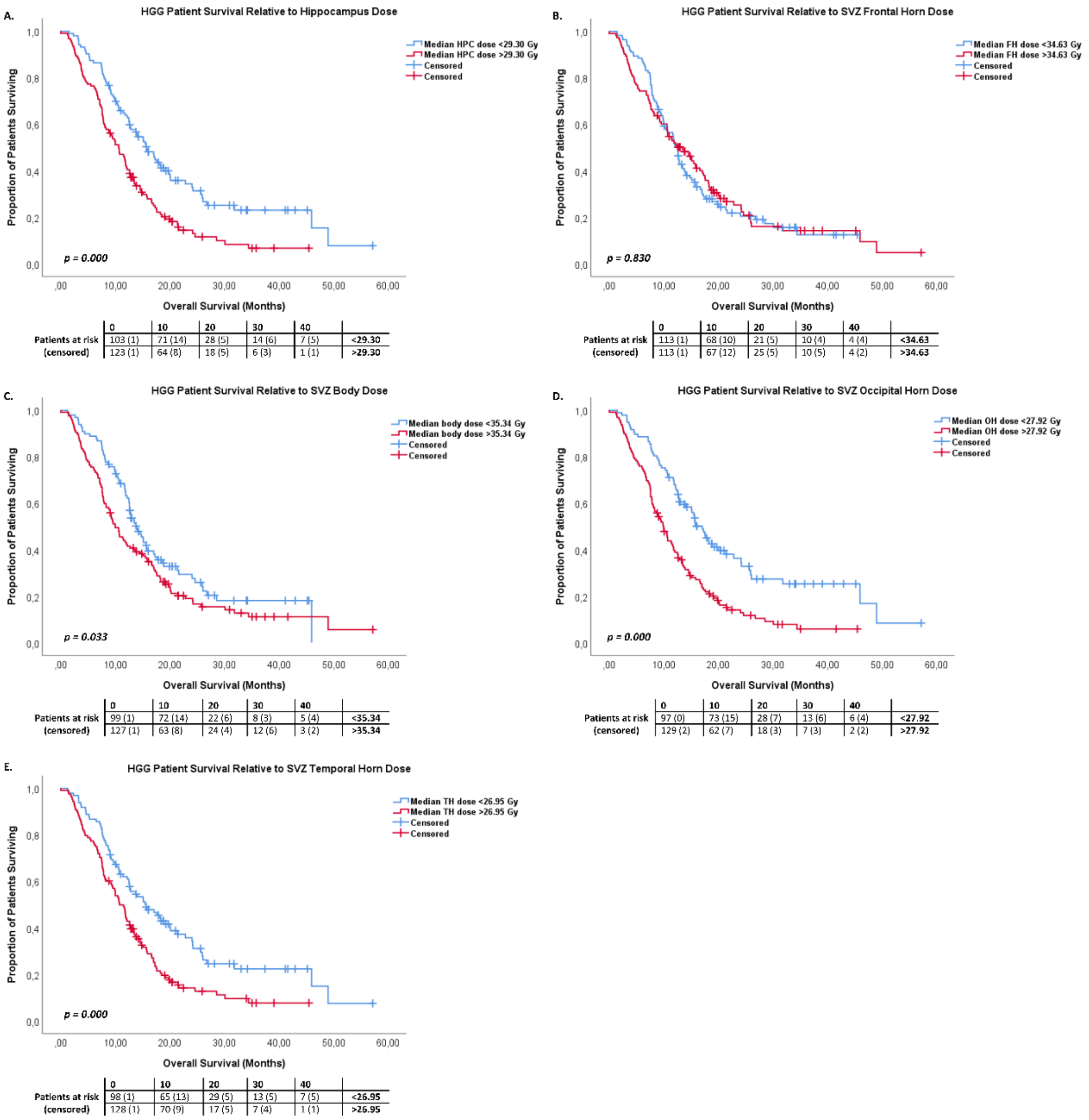


**Supplementary Figure 5.** Survival curves of different neurogenic structure doses in high-grade glioma (HGG) patients, that illustrate the difference in overall survival (OS) between high- and low-dose groups, stratified by the median of the mean doses received by each neurogenic structure. **A.** Survival curve of HGG patients relative to the median hippocampus dose (*p<0.001*). **B.** Survival curve of HGG patients relative to the median frontal horn dose (*p = 0.830*). **C.** Survival curve of HGG patients relative to the median body dose (*p = 0.033*). **D.** Survival curve of HGG patients relative to the median occipital horn dose (*p<0.001*). **E.** Survival curve of HGG patients relative to the median temporal horn dose (*p<0.001*).

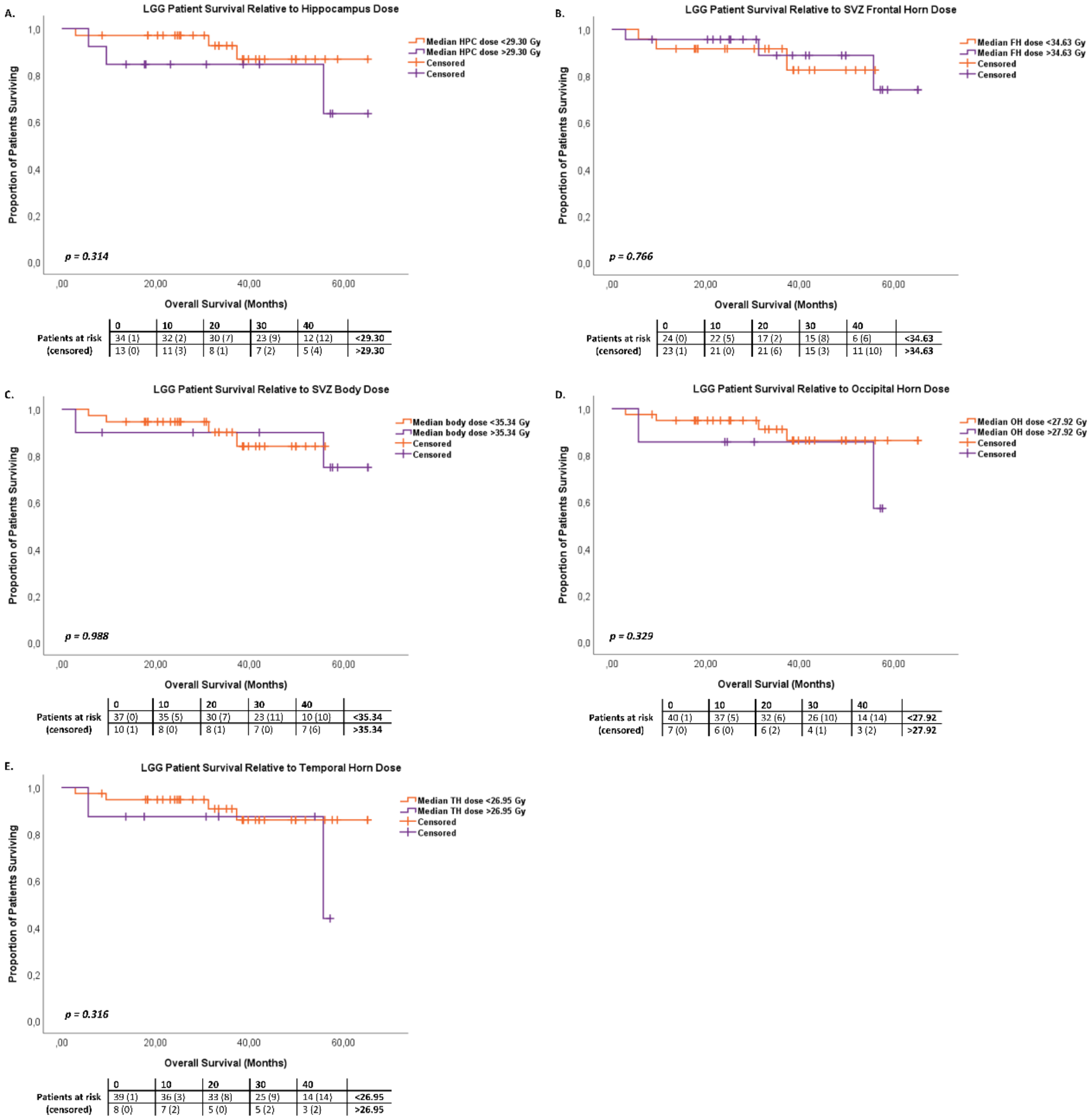


**Supplementary Figure 6.** Survival curves of different neurogenic structure doses in low-grade glioma (LGG) patients, that illustrate there is no difference in overall survival (OS) between high- and low-dose groups, stratified by the median of the mean doses received by each neurogenic structure. **A.** Survival curve of LGG patients relative to the median hippocampus dose (*p = 0.314*). **B.** Survival curve of LGG patients relative to the median frontal horn dose (*p = 0.766*). **C.** Survival curve of LGG patients relative to the median body dose (*p = 0.988*). **D.** Survival curve of LGG patients relative to the median occipital horn dose (*p = 0.329*). **E.** Survival curve of LGG patients relative to the median temporal horn dose (*p = 0.316*).

**Supplementary Table 3.** Cox regression analysis of ipsilateral and contralateral SVZ and SGZ doses on overall survival (OS) in high-grade glioma (HGG) patients and low-grade glioma (LGG) patients. Analysis is performed univariate for irradiation dose only, and multivariate corrected for different covariates.

| Patient diagnosis | Corrected for covariates | | | Ipsilateral SVZ | Contralateral SVZ | Ipsilateral SGZ | Contralateral SGZ |
|---|---|---|---|---|---|---|---|
| HGG | | | *p-value* | *<0.001* | *0.007* | *<0.001* | *0.001* |
| | | | HR | 1.030 | 1.019 | 1.018 | 1.016 |
| | | 95.0% CI for HR | Lower | 1.016 | 1.005 | 1.011 | 1.006 |
| | | | Upper | 1.045 | 1.032 | 1.025 | 1.025 |
| | Age, Sex, KPS, total intracranial volume | | *p-value* | *<0.001* | *0.002* | *<0.001* | *0.002* |
| | | | HR | 1.030 | 1.022 | 1.018 | 1.015 |
| | | 95.0% CI for HR | Lower | 1.015 | 1.008 | 1.010 | 1.005 |
| | | | Upper | 1.045 | 1.035 | 1.025 | 1.024 |
| | Age, Sex, KPS, total intracranial volume, MGMT, IDH, 1P/19Q deletion, Chemotherapy, Surgery-extent | | *p-value* | *0.005* | *0.021* | *<0.001* | *0.003* |
| | | | HR | 1.060 | 1.046 | 1.032 | 1.052 |
| | | 95.0% CI for HR | Lower | 1.017 | 1.007 | 1.015 | 1.017 |
| | | | Upper | 1.104 | 1.086 | 1.050 | 1.089 |
| LGG | | | *p-value* | *0.968* | *0.642* | *0.399* | *0.893* |
| | | | HR | 1.002 | 1.019 | 1.022 | 0.995 |
| | | 95.0% CI for HR | Lower | 0.927 | 0.941 | 0.971 | 0.918 |
| | | | Upper | 1.082 | 1.104 | 1.076 | 1.078 |
| | Age, Sex, KPS, total intracranial volume | | *p-value* | *0.562* | *0.814* | *0.626* | *0.686* |
| | | | HR | 0.969 | 1.010 | 1.018 | 0.979 |
| | | 95.0% CI for HR | Lower | 0.870 | 0.927 | 0.949 | 0.884 |
| | | | Upper | 1.078 | 1.101 | 1.091 | 0.085 |
| | Age, Sex, KPS, total intracranial volume, Chemotherapy, Surgery-extent | | *p-value* | *0.764* | *0.317* | *0.448* | *0.905* |
| | | | HR | 0.982 | 1.082 | 1.029 | 1.008 |
| | | 95.0% CI for HR | Lower | 0.869 | 0.927 | 0.955 | 0.888 |
| | | | Upper | 1.108 | 1.264 | 1.109 | 1.143 |

*MGMT* O6-methylguanine methyltransferase, *IDH* Isocitrate dehydrogenase, *HPC* Hippocampus, *SGZ* Subgranular zone, *SVZ* Subventricular zone, *HR* Hazard ratio of overall survival, *CI* Confidence interval.

**Supplementary Table 4.** Overview of the median ipsilateral and contralateral irradiation doses received by the subventricular zone (SVZ) and subgranular zone (SGZ) and Kaplan-Meier analysis of these doses on overall survival (OS), with median and mean estimates for HGG patients and mean estimates for LGG patients. Estimates are given in months.

| **Patient diagnosis** | | | **Ipsilateral SVZ** | | **Contralateral SVZ** | | **Ipsilateral SGZ** | | **Contralateral SGZ** | |
|---|---|---|---|---|---|---|---|---|---|---|
| | | **Median dose (Gy)** | 39.41 | | 20.45 | | 45.03 | | 12.54 | |
| | | **Dose group** | **High** | **Low** | **High** | **Low** | **High** | **Low** | **High** | **Low** |
| HGG patients | Median OS estimate (months) | | 11.5 | 13.8 | 10.7 | 14.1 | 10.7 | 15.5 | 10.6 | 15.8 |
| | 95,0% CI | Lower | 9.7 | 11.6 | 8.6 | 11.5 | 8.1 | 12.5 | 9.0 | 13.1 |
| | | Upper | 13.4 | 16.0 | 12.9 | 16.7 | 13.2 | 18.5 | 12.3 | 18.6 |
| | Mean OS estimate (months) | | 14.5 | 21.2 | 16.2 | 18.2 | 13.6 | 21.5 | 13.7 | 21.7 |
| | 95.0% CI | Lower | 12.2 | 17.1 | 13.1 | 15.5 | 11.3 | 17.7 | 11.5 | 17.7 |
| | | Upper | 16.7 | 25.3 | 19.3 | 20.9 | 15.9 | 25.3 | 15.8 | 25.6 |
| | | *p-value* | *0.035* | | *0.318* | | *0.002* | | *0.001* | |
| LGG patients | Mean OS estimate (months) | | 55.1 | 52.9 | 56.6 | 52.7 | 57.9 | 60.1 | 63.4 | 57.1 |
| | 95.0% CI | Lower | 43.9 | 48.9 | 47.1 | 48.1 | 50.3 | 53.3 | 60.0 | 49.9 |
| | | Upper | 66.3 | 57.0 | 66.1 | 57.2 | 65.6 | 66.9 | 66.7 | 64.4 |
| | | *p-value* | *0.722* | | *0.715* | | *0.792* | | *0.528* | |

*HPC* Hippocampus, *SGZ* Subgranular zone, *SVZ* Subventricular zone, *OS* Overall survival, *CI* Confidence interval.

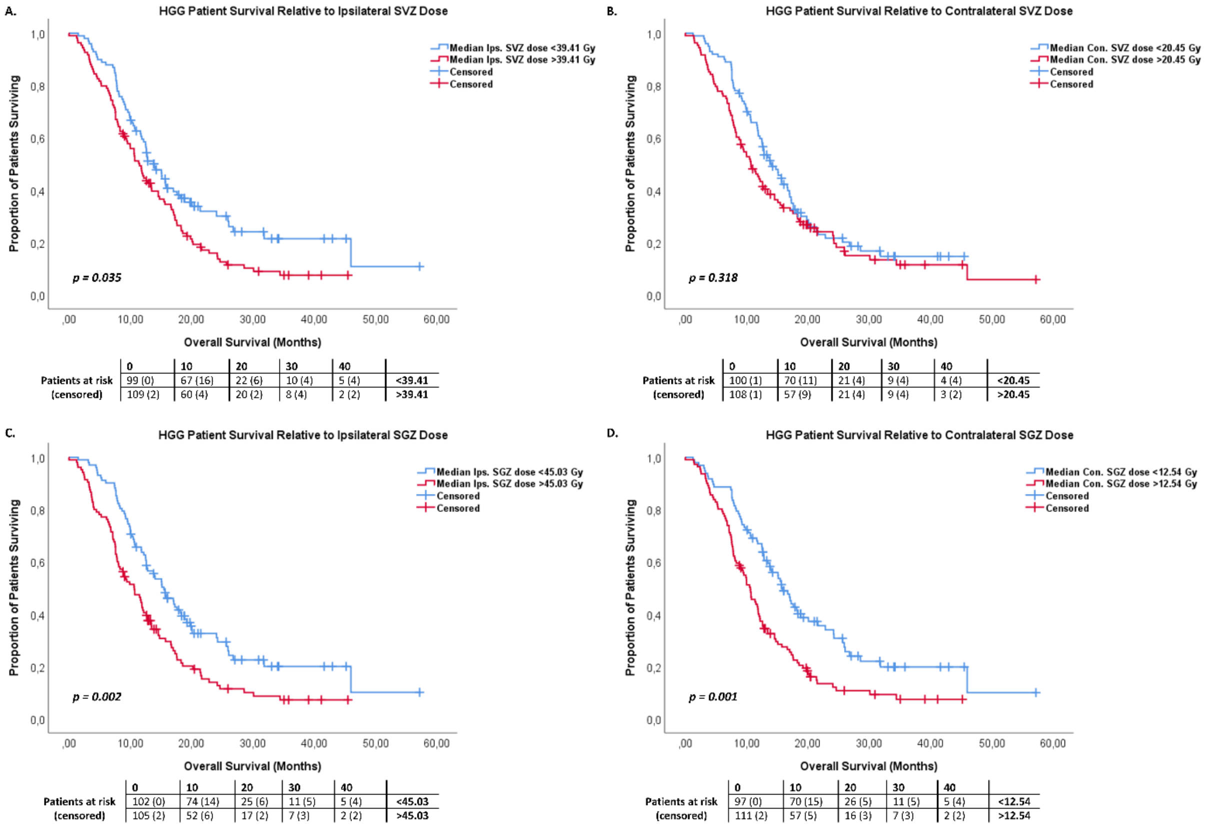


**Supplementary Figure 7.** Survival curves of ipsilateral and contralateral doses on the subventricular zone (SVZ) and subgranular zone (SGZ) in high-grade glioma (HGG) patients, that illustrate the difference in overall survival (OS) between high- and low-dose groups, stratified by the median of the mean ipsi- and contralateral doses on the SVZ and SGZ. **A.** Survival curve of HGG patients relative to the median ipsilateral SVZ dose (*p = 0.035*). **B.** Survival curve of HGG patients relative to the median contralateral SVZ dose (*p = 0.318*). **C.** Survival curve of HGG patients relative to the median ipsilateral SGZ dose (*p = 0.002*). **D.** Survival curve of HGG patients relative to the median contralateral SGZ dose (*p = 0.001*).

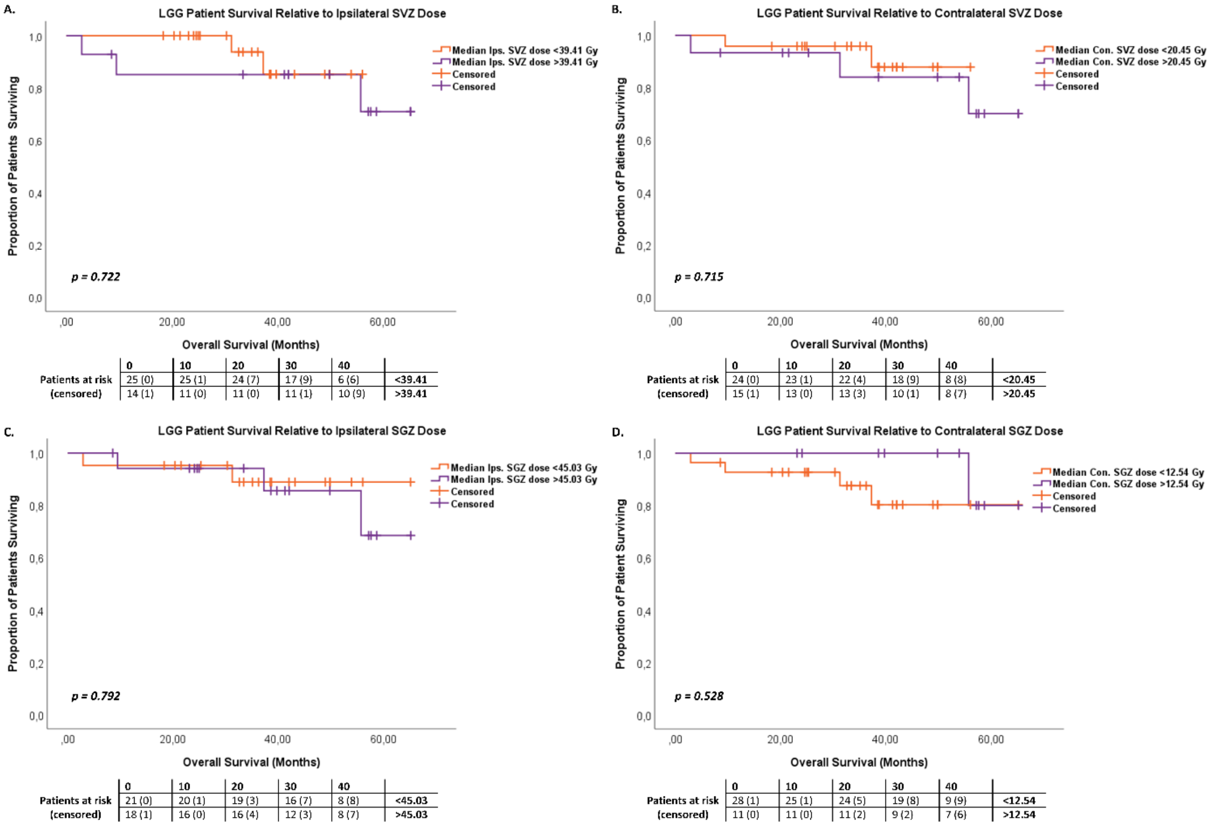


**Supplementary Figure 8.** Survival curves of ipsilateral and contralateral doses on the subventricular zone (SVZ) and subgranular zone (SGZ) in low-grade glioma (LGG) patients, that illustrate there is no difference in overall survival (OS) between high- and low-dose groups, stratified by the median of the mean ipsi- and contralateral doses on the SVZ and SGZ. **A.** Survival curve of LGG patients relative to the median ipsilateral SVZ dose ($p = 0.722$). **B.** Survival curve of LGG patients relative to the median contralateral SVZ dose ($p = 0.715$). **C.** Survival curve of LGG patients relative to the median ipsilateral SGZ dose ($p = 0.792$). **D.** Survival curve of HGG patients relative to the median contralateral SGZ dose ($p = 0.528$).

**References**


1 Greve, Douglas N. and Fischl, Bruce (2009) "Accurate and robust brain image alignment using boundary-based registration." *NeuroImage*, 48(1), pp. 63–72.

2 Jenkinson, Mark and Smith, Stephen (2001) "A global optimisation method for robust affine registration of brain images." *Medical Image Analysis*, 5(2), pp. 143–156.

3 Jenkinson, Mark, Bannister, Peter, Brady, Michael and Smith, Stephen (2002) "Improved Optimization for the Robust and Accurate Linear Registration and Motion Correction of Brain Images." *NeuroImage*, 17(2), pp. 825–841.

4 Radwan, Ahmed M., Emsell, Louise, Blommaert, Jeroen, Zhylka, Andrey, et al. (2021) "Virtual brain grafting: Enabling whole brain parcellation in the presence of large lesions." *NeuroImage*, 229, p. 117731.

5 Jenkinson, Mark, Beckmann, Christian F., Behrens, Timothy E.J., Woolrich, Mark W. and Smith, Stephen M. (2012) "FSL." *NeuroImage*, 62(2), pp. 782–790.

6 Fischl, Bruce (2012) "FreeSurfer." *NeuroImage*, 62(2), pp. 774–781.

7 Penny, William D, Friston, Karl J, Ashburner, John T, Kiebel, Stefan J and Nichols, Thomas E (2011) *Statistical parametric mapping: the analysis of functional brain images*, Elsevier.

8 Gaser, C, Hbm, R Dahnke - and 2016, undefined (n.d.) "CAT-a computational anatomy toolbox for the analysis of structural MRI data." *neuro.uni-jena.de*.

9 Manjó, José v, Coupé, Pierrick, Martí-Bonmatí, Luis, Collins, D Louis and Robles, Montserrat (2010) "Adaptive Non-Local Means Denoising of MR Images With Spatially Varying Noise Levels." *J. Magn. Reson. Imaging*, 31, pp. 192–203.

10 Ashburner, John and Friston, Karl J. (2005) "Unified segmentation." *NeuroImage*, 26(3), pp. 839–851.

11 Ashburner, John (2007) "A fast diffeomorphic image registration algorithm." *NeuroImage*, 38(1), pp. 95–113.

12 Tohka, Jussi, Zijdenbos, Alex and Evans, Alan (2004) "Fast and robust parameter estimation for statistical partial volume models in brain MRI." *NeuroImage*, 23(1), pp. 84–97.

13 Winterburn, Julie L., Pruessner, Jens C., Chavez, Sofia, Schira, Mark M., et al. (2013) "A novel in vivo atlas of human hippocampal subfields using high-resolution 3T magnetic resonance imaging." *NeuroImage*, 74, pp. 254–265.

14 Cherubini, Andrea, Spoletini, Ilaria, Péran, Patrice, Luccichenti, Giacomo, et al. (2010) "A multimodal MRI investigation of the subventricular zone in mild cognitive impairment and Alzheimer's disease patients." *Neuroscience letters*, 469(2), pp. 214–218.

15 Lee, Percy, Eppinga, Wietse, Lagerwaard, Frank, Cloughesy, Timothy, et al. (2013) “Evaluation of high ipsilateral subventricular zone radiation therapy dose in glioblastoma: a pooled analysis.” *International Journal of Radiation Oncology* Biology* Physics*, 86(4), pp. 609–615.

16 Chen, Linda, Guerrero-Cazares, Hugo, Ye, Xiaobu, Ford, Eric, et al. (2013) “Increased subventricular zone radiation dose correlates with survival in glioblastoma patients after gross total resection.” *International Journal of Radiation Oncology* Biology* Physics*, 86(4), pp. 616–622.

17 Hallaert, Giorgio, Pinson, Harry, van den Broecke, Caroline, Sweldens, Caroline, et al. (2021) “Survival impact of incidental subventricular zone irradiation in IDH-wildtype glioblastoma.” *Acta Oncologica*, 60(5), pp. 613–619.

18 Elicin, Olgun, Inac, Ebrar, Uzel, Esengul Kocak, Karacam, Songul and Uzel, Omer Erol (2014) “Relationship between survival and increased radiation dose to subventricular zone in glioblastoma is controversial.” *Journal of neuro-oncology*, 118(2), pp. 413–419.

19 Alvarez-Buylla, Arturo and Garcıa-Verdugo, Jose Manuel (2002) “Neurogenesis in adult subventricular zone.” *Journal of Neuroscience*, 22(3), pp. 629–634.

20 Rhoton Jr., Albert L (2002) “The Lateral and Third Ventricles.” *Neurosurgery*, 51(suppl_4), pp. S1-207-S1-271.
